# Supraspinal control of motoneurons after paralysis enabled by spinal cord stimulation

**DOI:** 10.1101/2023.11.29.23298779

**Authors:** Josep-Maria Balaguer, Genis Prat-Ortega, Nikhil Verma, Prakarsh Yadav, Erynn Sorensen, Roberto de Freitas, Scott Ensel, Luigi Borda, Serena Donadio, Lucy Liang, Jonathan Ho, Arianna Damiani, Erinn Grigsby, Daryl P. Fields, Jorge A. Gonzalez-Martinez, Peter C. Gerszten, Lee E. Fisher, Douglas J. Weber, Elvira Pirondini, Marco Capogrosso

**Author notes:** Co-first authors (Josep-Maria Balaguer, Genis Prat-Ortega).

## Abstract

Spinal cord stimulation (SCS) restores motor control after spinal cord injury (SCI) and stroke. This evidence led to the hypothesis that SCS facilitates residual supraspinal inputs to spinal motoneurons. Instead, here we show that SCS does not facilitate residual supraspinal inputs but directly triggers motoneurons action potentials. However, supraspinal inputs can shape SCS-mediated activity, mimicking volitional control of motoneuron firing. Specifically, by combining simulations, intraspinal electrophysiology in monkeys and single motor unit recordings in humans with motor paralysis, we found that residual supraspinal inputs transform subthreshold SCS-induced excitatory postsynaptic potentials into suprathreshold events. We then demonstrated that only a restricted set of stimulation parameters enables volitional control of motoneuron firing and that lesion severity further restricts the set of effective parameters. Our results explain the facilitation of voluntary motor control during SCS while predicting the limitations of this neurotechnology in cases of severe loss of supraspinal axons.

## INTRODUCTION

In the last two decades, a series of animal and clinical studies showed that epidural electrical spinal cord stimulation (SCS) can restore some degree of motor control of lower and upper limbs in people with spinal cord injury and stroke^1–7^. The most striking observation that emerged from clinical trials is the evidence that, during SCS, humans with motor paralysis appear able to voluntarily control their previously paralyzed limbs^2,8^. In other words, SCS does not generate movement per se, but rather facilitates volitional movement. In an effort to explain these results, mechanistic investigations on SCS focused either on the identification of the neural pathways that are directly recruited by SCS^9–12^ or the involvement of movement-generating spinal circuits such as reflexes^10–12^, synergies^13^ and central pattern generators^14–16^. However, these studies never considered the contribution of residual supraspinal inputs, thereby failing to explain the facilitation of voluntary movements. This poses a problem because the current design of SCS protocols is focused on the recruitment of spinal neural structures^5,11,17^, without considering the role of volitional inputs. Understanding how volitional inputs can shape motor output during SCS could lead to more effective stimulation protocols targeted to maximize residual volitional input.

One of the most popular views into how SCS produces this facilitation is that electrical stimulation exerts a neuromodulatory effect on spinal circuits^2,8,18^. More specifically, SCS would increase the excitability of spinal circuits and, particularly, spinal motoneurons, making them more responsive to volitional residual supraspinal inputs^2,8^. In other words, through this neuromodulatory mechanism, SCS would enable descending fibers that survive the injury to regain control of motoneurons below the lesion. Unfortunately, this conjecture only provides a phenomenological description of the effects of SCS but fails to propose an experimentally verifiable mechanism of action by which SCS restores motor function.

In order to transform this conjecture into an experimentally verifiable hypothesis, we started from identified properties and mechanisms of SCS. It is well-established that SCS directly activates sensory afferents in the dorsal roots and dorsal columns^9–11,19,20^. Importantly, these afferents form mono- and polysynaptic excitatory connections to spinal motoneurons and convey SCS-mediated inputs to motoneurons. Thus, stimulation of the sensory afferents could provide motoneurons with additional excitatory drive after paralysis. Indeed, studies in monkeys showed that SCS pulses potentiate monosynaptic corticospinal motor-evoked potentials^21^. Here, we started from this body of electrophysiological evidence to study the effects of excitatory sensory inputs elicited by SCS on volitional control of spinal motoneurons.

We built on established biophysics of motoneuron membrane dynamics^22–25^ to calculate their membrane potential while receiving SCS-mediated excitatory drive from sensory afferents and excitatory inputs from residual supraspinal fibers. We found that motoneurons can fire during voluntary drive by transforming subthreshold sensory inputs generated by SCS pulses into suprathreshold events, thereby mimicking voluntary activity. However, the SCS parameter space at which this phenomenon occurred shrunk as a function of lesion severity. We validated these predictions with electrophysiology in two monkeys, two persons with mild stroke and one person with severe spinal cord injury, who were implanted with SCS leads. Our results suggest that SCS does not facilitate voluntary drive. Instead, SCS directly generates motoneuron action potentials, but only when residual supraspinal drive is present.

## RESULTS

### Fundamental representation of the biophysics of SCS

To identify the membrane mechanisms underlying motoneuron recruitment during SCS (**Fig. 1a**), we started from four assumptions grounded on experimental evidence (**Fig. 1b**). First, SCS directly recruits large-diameter sensory afferents, particularly, Ia afferents^10^. Second, electrical stimulation causes simultaneous depolarization in all recruited afferents, thereby producing non-natural volleys of action potentials. Third, in contrast to this non-natural input, supraspinal drive can be modeled as a stochastic process of non-synchronized axon firing^26,27^. Fourth, a lesion reduces the number of available excitatory supraspinal fibers, thus impacting the ability of the cortex to produce action potentials in spinal motoneurons.

**Fig. 1.**
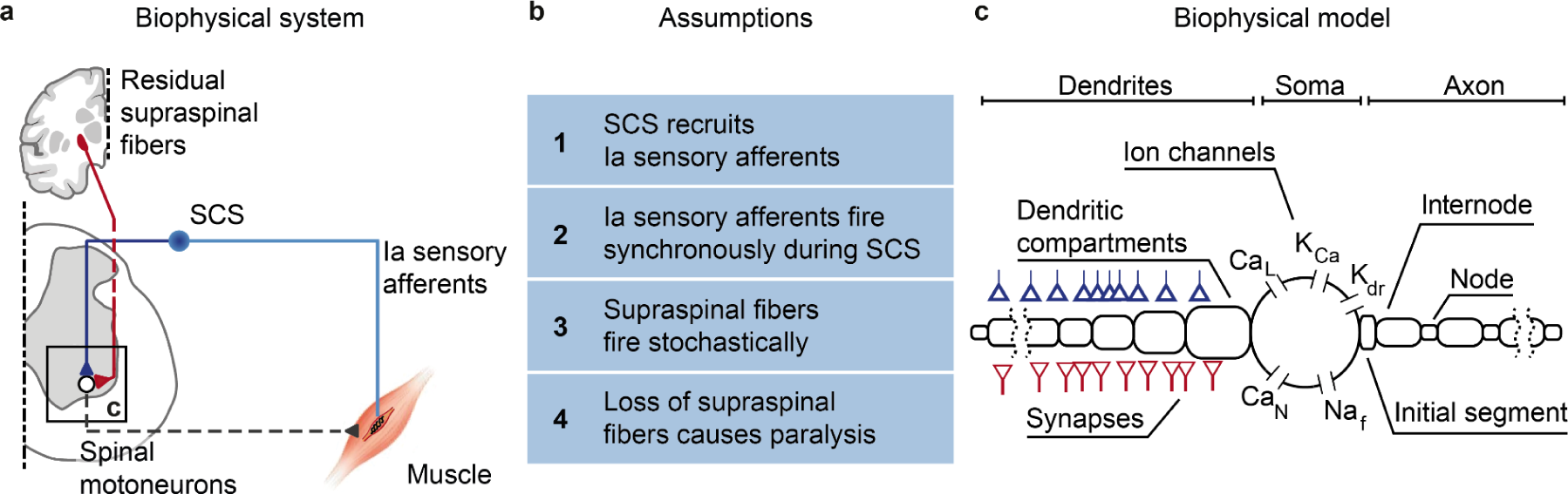
Biophysical model with underlying assumptions. **a**, Schematic of the spinal circuit involved in the recovery of voluntary movement during SCS composed of the monosynaptic Ia-to-motoneuron reflex circuit and supraspinal monosynaptic connections. Spinal motoneurons receive excitatory inputs from residual supraspinal fibers and Ia afferents recruited by SCS. **b**, Four assumptions grounded on experimental evidence that constitute the biophysical model of spinal motoneurons. **c**, Three-element biophysical model: motoneuron, Ia afferents inputs and supraspinal inputs. Biophysical model of spinal alpha motoneuron encompasses an electronic-equivalent dendritic tree, multicompartment soma and myelinated axon, connected through an initial axon segment. Motoneuron membrane dynamics follow dedicated Hodgkin–Huxley equations for multiple ion channels: L-type Ca2+, Ca2+-activated K, delayed rectifier K+, N-type Ca2+, and nonlinear fast Na+.

We then designed the simplest known circuit involved in voluntary motor control: motoneurons, Ia afferents and supraspinal fibers (**Fig. 1c**). In this three-element model, the membrane potential of spinal motoneurons is described with a modified Hodgkin-Huxley biophysical representation^25^. Instead, Ia afferents and supraspinal fibers are treated as point-processes with given firing rates connected to synaptic boutons, triggering excitatory postsynaptic potentials (EPSPs) in the motoneurons.

We modeled the stimulation intensity as the number of Ia afferents activated by SCS, allowing us to vary between no activation at all and recruitment of the entire Ia population^24^. Recruited Ia afferents elicit synchronous volleys that fire at a given SCS frequency, a phenomenon that holds for stimulation frequencies up to 500 Hz^24^. Alternatively, voluntary commands are conceptualized by time-varying firing rates in the supraspinal Poisson inputs. Variability is introduced at every element of this circuit (see methods, **Extended Data Fig. 1a-c**). Finally, we modeled a lesioned system by modifying ion conductances of the spinal motoneurons membrane (see methods) and by reducing the number of available supraspinal fibers, thereby simulating different degrees of lesion severities.

With this model, we calculated the changes in membrane potentials of spinal motoneurons caused by simultaneous supraspinal and SCS inputs. We then used it to explore the effects of SCS parameters on the ability of residual supraspinal inputs to produce firing rates in spinal motoneurons, i.e., the ability to recover voluntary motor control.

### Supraspinal inputs transform subthreshold SCS-mediated EPSPs into action potentials

Using our model, we inspected the membrane potential of motoneurons before and after a lesion. Prior to a lesion, supraspinal inputs generated regular and sustained action potentials in the motoneuron membrane (**Fig. 2a**, top and middle panels). In our model, the slow calcium dynamics of spinal motoneurons expectedly limited their firing rate to 40 Hz^27^ (**Fig. 2a**, bottom panel). We then represented a lesion by suppressing the activity of about 70% of supraspinal fibers (**Fig. 2b**, white left). Incoming EPSPs produced a mean tonic depolarization in the membrane of 5.6 mV, ramping up within 100 ms after the start of the simulated supraspinal activity. This tonic depolarization effectively set the motoneuron membrane closer to firing threshold (around -60 mV). However, it was insufficient to produce action potentials, thus leading to simulated paralysis. We then simulated a train of subthreshold SCS that alone (i.e., without supraspinal inputs) did not suffice to directly produce action potentials in motoneurons (**Fig. 2b**, blue middle). As opposed to the tonic membrane depolarization produced by residual natural inputs, subthreshold SCS induced a transient depolarization. This transient depolarization occurred through discrete EPSPs events in consequence of the synchronized recruitment of Ia afferents following each stimulation pulse.

**Fig. 2.**
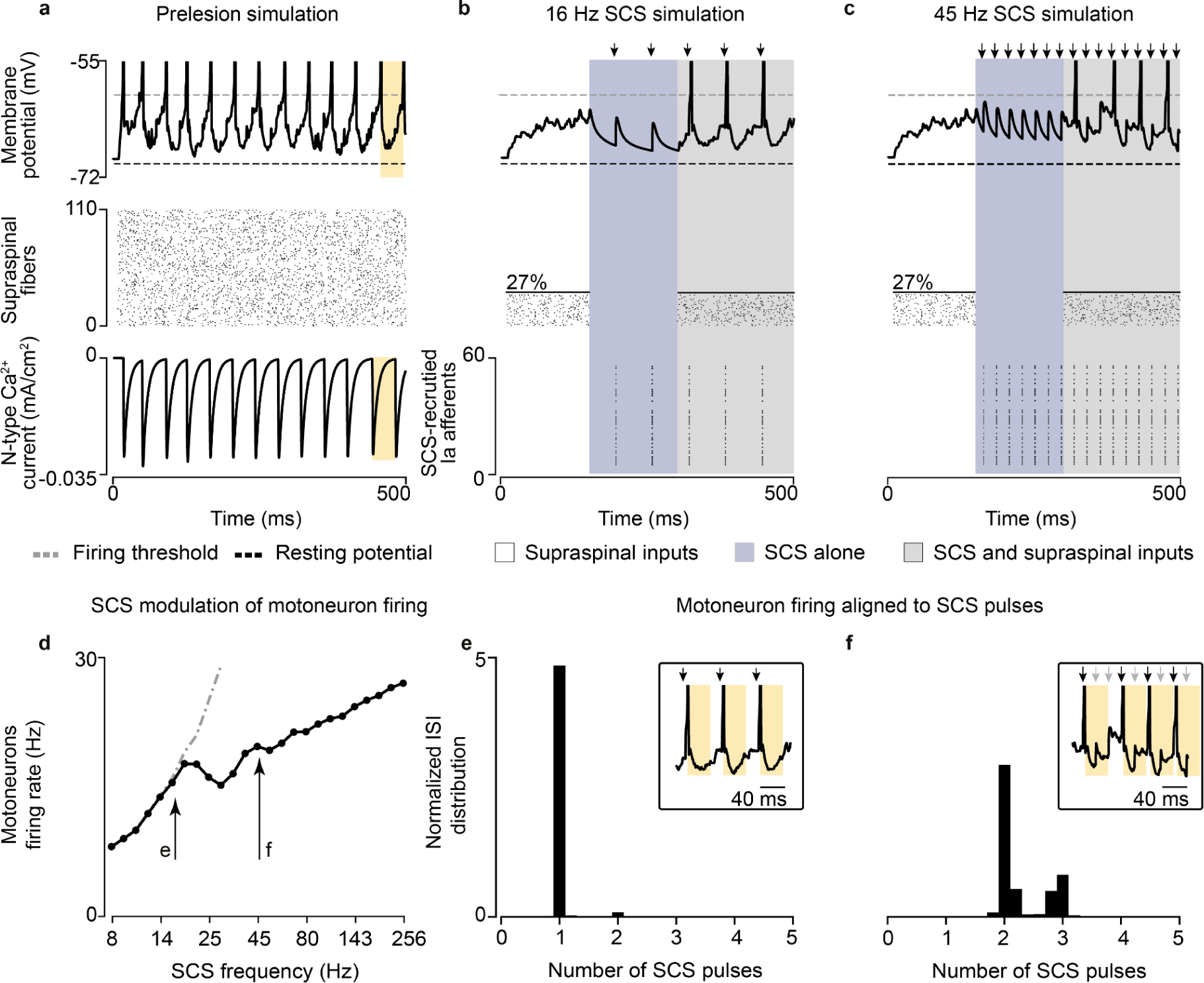
Supraspinal input facilitates SCS. **a,** Simulation of the membrane potential of a single motoneuron driven by supraspinal fibers prior to the lesion (top), raster plot of each supraspinal fiber firing stochastically (middle), ionic current of N-type Ca2+ of the motoneuron (bottom). Motoneuron firing rate is limited by the slow calcium dynamics (yellow). **b**, Simulation of the membrane potential of a single motoneuron after the lesion (white background), during SCS alone (16 Hz, 50% amplitude; blue) and concurrent residual supraspinal inputs and SCS (gray, top), raster plot of each residual supraspinal fiber firing stochastically after the lesion (middle), raster plot of each Ia afferent firing synchronously after each SCS pulse (bottom). **c**, Same as **b** at a higher SCS frequency (45 Hz, 50% amplitude). **d**, Mean motoneuron pool firing rate as a function of SCS frequencies, coupled up to 19 Hz (i.e., motoneuron firing rate equals stimulation frequency up to 19 Hz SCS represented by the dashed gray line) and effectively decoupled at higher stimulation frequencies while still increasing motoneuron firing. **e**, Probability distribution of the motoneuron pool ISI normalized by the stimulation interpulse interval (16 Hz SCS), indicating the number of SCS pulses between action potentials (in this case, 1 SCS pulse). Inset indicates the membrane potential of a motoneuron during concurrent supraspinal inputs and SCS, highlighting in yellow the time constraint of the slow calcium dynamics. **f**, Same as **e** at a higher SCS frequency (45 Hz). Arrows indicate stimulation pulses. Light arrows indicate skipped pulses elicited within the slow calcium dynamics in inset in **f**.

Next, we combined subthreshold SCS with residual supraspinal inputs, which led to a robust production of action potentials that seemed aligned to the stimulation pulses (**Fig. 2b**, gray right). Indeed, when calculating the interspike interval (ISI) of the motoneuron pool, we found that it coincided with the stimulation interpulse interval (**Fig. 2e**). In other words, for each stimulation pulse, motoneurons produced an action potential. Subsequently, we increased the stimulation frequency to further explore this phenomenon and found that, for SCS frequencies above 19 Hz (still subthreshold), motoneurons were not able to generate action potentials for each SCS pulse because of the time constraint in their slow calcium dynamics (**Fig. 2c,f; Extended Data Fig. 1d Extended Data Fig. 2**). Specifically, motoneurons membrane skipped those SCS pulses that were elicited within the calcium dynamics following a motoneuron action potential (**Fig. 2f** light arrows in the inset). In turn, the number of skipped pulses increased as a function of the SCS frequency (**Extended Data Fig. 3c-g)**. However, action potentials still occurred only in consequence of SCS pulses and, despite this decoupling above 19 Hz SCS, higher stimulation frequencies increased non-monotonically motoneuron pool firing rates (**Fig. 2d)**.

In summary, we observed that tonic depolarization from residual supraspinal inputs transforms subthreshold SCS-mediated transient depolarizations into suprathreshold events in the motoneuron membrane. Yet, motoneurons were not able to follow these suprathreshold inputs for all frequencies, skipping some stimulation pulses before producing action potentials. This resulted in characteristic peaks in motoneuron ISI distributions at integers of SCS interpulse intervals.

### Voluntary vs involuntary motoneuron activity during SCS

Given the influence of SCS frequency on motoneuron firing rates, we extended our analysis to study the effects of SCS parameters on the production of volitional motoneuron activity. To this end, we quantified the firing activity in a simulated pool of motoneurons during SCS alone (**Fig. 3a**) and during concurrent SCS and residual input (**Fig. 3b**), while varying SCS amplitude (i.e., percentage of Ia afferents recruited, 10-100%) and frequency (8-256 Hz). We found that, for certain parameters (a nonlinear trade-off above 30-70% and 12-124 Hz), SCS alone directly produced sustained motoneuron firing rate (>8 Hz)^28^, even without supraspinal input (**Fig. 3a**). This well-known phenomenon has been thoroughly described in animal and human studies as suprathreshold SCS^3,17,29^. This combination of parameters sharply differed from the subthreshold parameters (below 30-70% and 12-124 Hz, **Fig. 3a**), where SCS alone did not produce firing rates in the motoneuron pool, being effectively “sub-threshold”^8^.

**Fig. 3.**
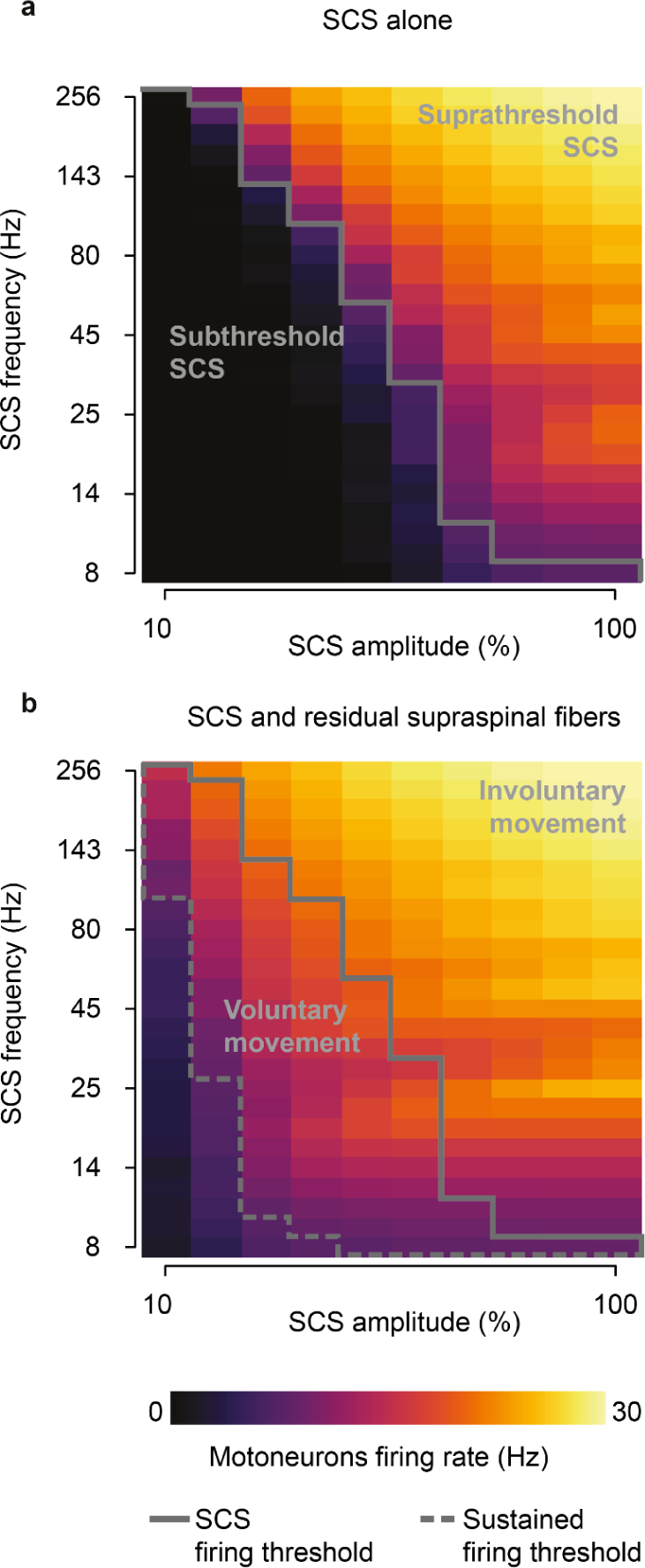
SCS parameter regimes. **a,** Simulated heatmap of mean firing rates of a motoneuron pool during SCS alone (5 second simulation) for a range of frequencies and amplitudes. Solid line separates subthreshold from suprathreshold SCS parameters. **b,** Same as **a** during concurrent SCS and residual supraspinal inputs innervating the pool of motoneurons. Dashed line separates the combinations of parameters that produce sustained motoneuron firing rates (>8 Hz). The parameters between the grey lines represent the combination of SCS parameters that produce sustained firing rate only with concurrent SCS and residual supraspinal inputs (i.e., voluntary movement regime).

When we combined SCS with residual supraspinal inputs, we observed a general increase in motoneuron firing rates (**Fig. 3b**). Interestingly, most of the subthreshold SCS parameters that led to sparse or no firing rate showed sustained production of motoneuron firing rates. This pattern presented two large “regimes’’ in the SCS-parameter space that we termed *voluntary* and *involuntary movement regimes* (**Fig. 3b; Extended Data Fig. 4**). In the voluntary movement regime, SCS-mediated EPSPs became suprathreshold (only with concurrent inputs from residual supraspinal fibers) and motoneurons showed sustained firing rates (8-20 Hz). Instead, in the involuntary movement regime, SCS dominated motoneuron firing rates (20-30 Hz), with little influence from incoming residual supraspinal inputs.

In summary, we showed that there exists a well-defined range of stimulation parameters in which residual supraspinal inputs transform subthreshold SCS pulses into suprathreshold events.

### Residual supraspinal input can modulate motoneuron firing rate

The fact that residual supraspinal inputs can transform subthreshold SCS pulses into action potentials could imply that residual supraspinal inputs solely act as a gating mechanism. In this case, motoneuron firing rates would be fully controlled by SCS parameters and volitional modulation of forces would be impossible. Since this would contradict experimental evidence in humans, we explored the role of residual supraspinal inputs in the firing rates of motoneurons during SCS. We simulated motoneuron output during 45 Hz SCS for different strengths of residual supraspinal inputs by changing supraspinal mean firing. We observed that the motoneuron pool firing rates linearly increased with residual supraspinal fibers firing rates during SCS amplitude within the voluntary regime (i.e., 731.7% modulation of the motoneuron firing rate at 40% SCS amplitude). Instead, in the involuntary regime, motoneuron firing rates were not affected by residual supraspinal inputs (i.e., 11.5% modulation at 80% SCS amplitude, **Fig 4a**). We hypothesized that this modulation could occur by two mechanisms: 1) controlling the number of motoneurons that respond to SCS and 2) controlling the number of skipped pulses by recruited motoneurons.

**Fig. 4.**
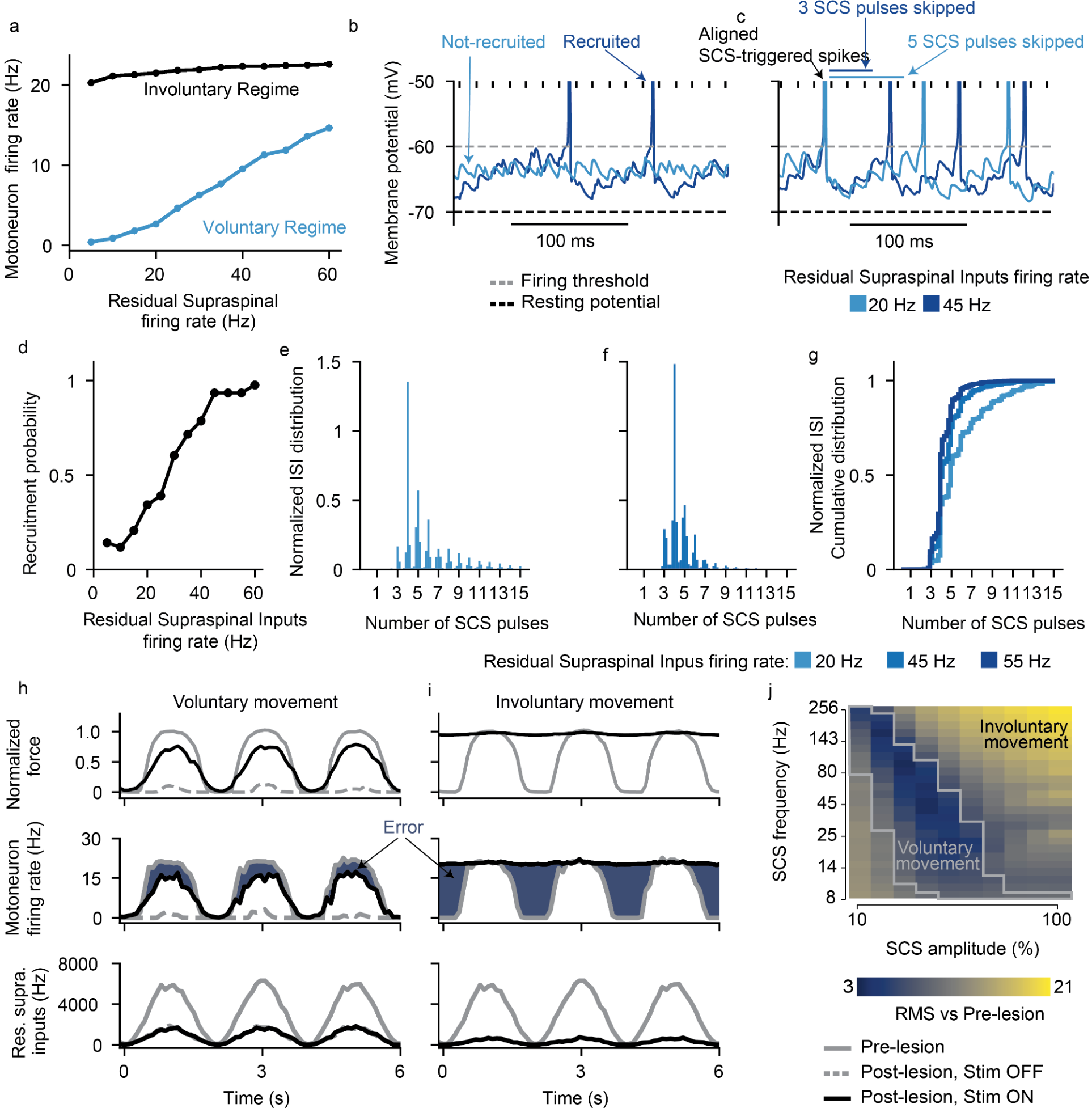
Residual supraspinal inputs can modulate motoneuron firing rate during SCS. **a**, Motoneuron pool firing rate (5 seconds simulations) as a function of residual supraspinal inputs firing rate in the voluntary movement regime (SCS parameters: 45 Hz and 30% amplitude) and involuntary regime (SCS parameters: 45 Hz and 80% amplitude). **b**, Membrane potential of the same motoneuron with residual supraspinal inputs at 20 and 45 Hz. This motoneuron is recruited at 45 Hz but not at 20 Hz. **c** Membrane potential of another motoneuron that skips 3 SCS pulses at 20 Hz and 4 or 5 at 45 Hz. SCS parameters: 69 Hz and 30% amplitude. **d,** Percentage of motoneurons recruited (with a firing rate >8 Hz) as a function of the residual supraspinal fibers firing rate **e,f**, Two examples of ISI normalized probability distributions for residual supraspinal firing rates at 20 and 45 Hz during SCS (69 Hz, 30%). **g,** ISI normalized cumulative distributions show that residual supraspinal inputs can modulate motoneuron firing rate by controlling the number of skipped SCS pulses. **h-i,** Fine movement task in the voluntary (69 Hz, 30% amplitude) and involuntary (69 Hz, 80% amplitude) movement regimes. Firing rates are computed in 100 ms bin windows. From bottom to top: supraspinal inputs that follow a sinusoidal wave, motoneuron firing rate and normalized force. Blue area indicates the difference in firing rate from the prelesion case (i.e., error). **j,** Root-mean square (RMS) of the difference between the motoneuron firing rate pre- vs postlesion (dark blue regions in **h** and **i**) for all simulated SCS parameters. The gray lines in the heatmap indicate the voluntary movement regime defined in the isometric task (**Fig. 3b**).

We inspected simulated membrane behavior to test for the first mechanism and found that, if residual supraspinal inputs was sufficiently strong (i.e., higher residual supraspinal fibers firing rates), it could induce stronger depolarization of motoneurons membrane, thereby bringing motoneurons closer to firing threshold and increasing the number of recruited motoneurons (see an example of a motoneuron that was recruited by SCS with high residual supraspinal inputs (dark blue), but not low (light blue), **Fig. 4b**). Extending these results over the motoneuron pool, the number of motoneurons that responded to SCS increased with higher residual supraspinal fibers firing rates (i.e., recruitment probability increase of 82.7% from supraspinal fibers firing at 5 Hz vs 60 Hz, **Fig. 4d**). This means that residual supraspinal inputs can actively control the number of motoneurons that respond to SCS.

Second, we found that, as the firing rate of the residual supraspinal fibers increased, motoneuron depolarization was not only stronger but brought motoneurons to threshold faster. This increase in depolarization speed during continuous SCS pulses effectively decreased the number of SCS pulses skipped (i.e., SCS pulses that were not transformed into suprathreshold events). For instance, the motoneuron in **Fig 4c** skipped 4 to 5 SCS pulses with residual supraspinal fibers firing rate of 20 Hz, but it only skipped 3 with a residual supraspinal fibers firing rate of 45 Hz. This means that the residual supraspinal inputs can also control the number of skipped SCS pulses for motoneurons that respond to SCS. Indeed, ISI histograms normalized by the SCS interpulse interval were shifted towards smaller values for stronger residual supraspinal inputs (**Fig. 4e-g**).

To illustrate the impact of SCS on fine motor control, we used a task where residual supraspinal inputs followed a sinusoidal function and we estimated the force produced by a pool of motoneurons^30^ (see methods, **Fig. 4h,i**). We estimated the error by comparing the firing rate of the pool of motoneurons with the prelesion case. (**Fig. 4j**), thereby assessing SCS parameters efficacy. We found that SCS parameters in the voluntary movement regime assisted residual supraspinal inputs to follow the task (**Fig. 4h,j**). Instead, SCS parameters in the involuntary movement regime impeded residual supraspinal inputs modulation of motoneuron activity, yielding high task errors (**Fig. 4i,j**).

In summary, our biophysical model predicted that, while every action potentials is triggered by an SCS pulse, residual supraspinal inputs can still volitionally modulate motoneuron activity by changing the number of motoneurons that respond to SCS and the number of SCS pulses that they skip effectively, affecting recruitment and firing rates of motoneurons.

### Experimental validation in monkeys

We conducted animal experiments to validate the underlying model assumptions and predictions. We designed an experimental setup in anesthetized macaque monkeys (n=2; Mk-JC, Mk-Ka), who share with humans monosynaptic corticospinal innervations of spinal motoneurons^31,32^. Briefly, to replicate our simulations in a controlled fashion, we applied SCS at C6-C8 cervical spinal segments, while manipulating supraspinal input by means of deep-brain stimulation of the internal capsule^33^, which is a deep-brain structure hosting corticospinal fibers. In parallel, we measured motoneuron output directly, via intraspinal neural recordings, and indirectly, via evoked intramuscular electromyographic (EMG) in the hand muscles (**Fig. 5a**).

**Fig. 5.**
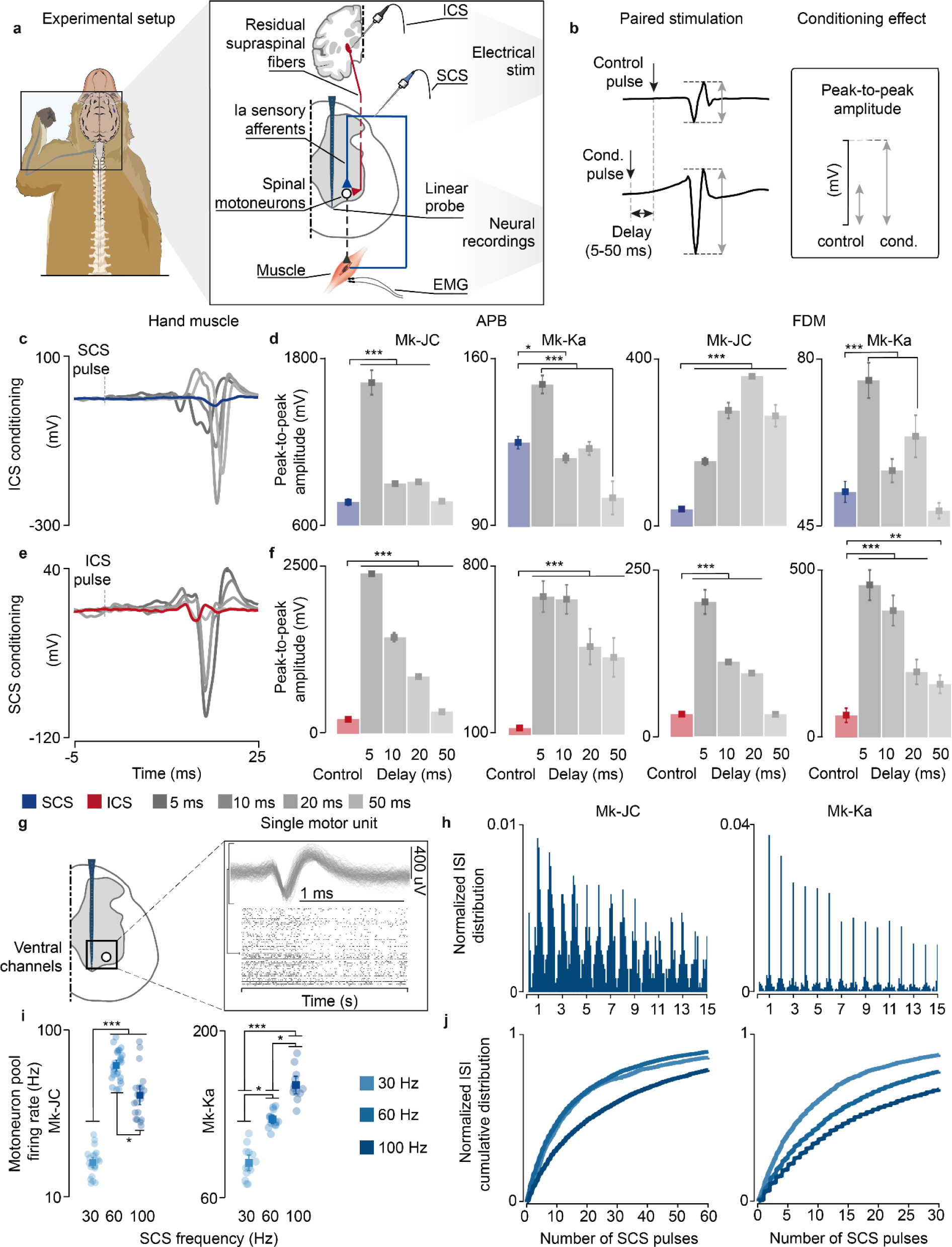
Experimental validation of model assumptions and predictions in monkeys. **a,** Experimental setup in anesthetized monkeys and schematic of the spinal circuits involved during dorsal root stimulation and activation of corticospinal fibers via deep-brain stimulation of the internal capsule. In response to the stimulation, EMG hand muscles responses were recorded and intraspinal spikes captured. **b**, Schematic of the paired stimulation protocol. A single pulse of SCS (top) is facilitated with ICS conditioning at different delays (bottom) and vice versa, yielding greater peak-to-peak amplitudes of the EMG traces of the conditioned pulse. **c**, Stimulation-triggered average of the EMG responses when conditioning with ICS a single pulse of SCS at different delays in FDM muscle in Mk-JC. **d**, Peak-to-peak amplitudes of the EMG responses in the hand muscles (FDM, APB) conditioned with ICS in both monkeys. **e**, Same as **c** when conditioning with SCS a single pulse of ICS. **f**, Same as **d** conditioning with SCS. Statistical quantification of the peak-to-peak amplitudes (***p<0.001; **p<0.01; *p<0.05; Kruskal-Wallis test with 117, 116, 119, 113, 121 points for FDM muscle for control, 5 ms, 10 ms, 50 ms, 100 ms delay during ICS conditioning in Mk-JC; 118, 96, 113, 100, 119 points for APB muscle during ICS conditioning in Mk-JC; 58, 64, 61, 53, 58 points for FDM muscle during iCS conditioning in Mk-Ka; 71, 60, 60, 49, 61 points for APB muscle during iCS conditioning in Mk-Ka. 118, 119, 116, 118, 117 points for FDM muscle for control, 5 ms, 10 ms, 50 ms, 100 ms delay during SCS conditioning in Mk-JC; 117, 97, 119, 98, 114 points for APB muscle during SCS conditioning in Mk-JC; 59, 60, 59, 67, 58 points for FDM muscle during SCS conditioning in Mk-Ka; 58, 60, 60, 71, 58 for APB muscle during SCS conditioning in Mk-Ka). **g**, Samples of a raster plot corresponding to sorted motor unit waveforms recorded from the ventral channels of the linear probe (motor units sorted from a 30-second trial). **h**, Histograms of normalized ISI probability distribution of the sorted motor units during 100 Hz SCS in both monkeys. **i**, Firing rates of the sorted motor units for different frequencies in Mk-JC (left) and Mk-Ka (right). Statistical quantification of the firing rates (***p<0.001; **p<0.01; *p<0.05; Kruskal-Wallis test with 22, 26 and 21 points for 30 Hz (0.14 mA), 60, Hz (0.14 mA) and 100 Hz (0.11 mA) in Mk-JC. 14, 13 and 12 points for in Mk-Ka (0.7 mA SCS). **j**, Normalized ISI cumulative distribution of the sorted motor units during continuous subthreshold SCS at 30, 60 and 100 Hz. Square and error bars indicate the mean distribution of the data with 95% confidence interval.

Our main model prediction was that supraspinal fibers can modulate motoneuron membrane and reduce their threshold for incoming SCS-mediated EPSPs. If this is true, conditioning a pulse of SCS with a single pulse of internal capsule stimulation (ICS) should facilitate SCS and produce larger SCS-triggered spinal reflexes in targeted muscles. Moreover, this facilitation should occur at latencies that are compatible with the time coincidence of monosynaptic Ia-afferent mediated EPSPs and cortex-mediated excitatory inputs^21^.

We thus designed a stimulation protocol to condition SCS with a pulse of ICS at different delays (5-50 ms, **Fig. 5b**). In both monkeys, mean peak-to-peak of SCS-induced reflex amplitudes significantly increased when SCS was conditioned by a pulse of ICS, demonstrating that corticospinal inputs can facilitate SCS reflexes (Mk-JC: 111.3%, 17.2%, 18.9%, 7.2% in APB; 285.8%, 590.9%, 793.3%, 557.7% in FDM. Mk-Ka: 19.6%, -5.12%, -1.8%, -18.3% in FDM; 44.6%, 8.5%, 22.4%, -7.7% in APB, **Fig. 5c,d**). Specifically, the greatest reflex peak-to-peak amplitude was obtained at 5 ms delay in Mk-Ka, when both EPSPs coincided in the hand muscles (**Extended Data Fig. 5a**). Another important feature of our model is that both Ia afferents and supraspinal fibers excite motoneurons through fast glutamatergic synapses, resulting in a similar effect on the motoneuron membrane. Thus, SCS-induced EPSPs should modulate corticospinal motor-evoked potentials. Therefore, we repeated the same experiments but now conditioning ICS with a pulse of SCS at different delays. Indeed, corticospinal motor-evoked potentials mean peak-to-peak amplitudes significantly increased in comparison to control (i.e., SCS alone) in both monkeys, in contrast to previous reported results^21^ (**Fig. 5e,f**). These results show that both inputs can depolarize membrane potentials of motoneurons and facilitate a subsequent EPSP.

Second, we validated the assumption that SCS recruits sensory afferents eliciting synchronous volleys at a given stimulation frequency (**Fig. 1b**). To do so, we inspected postsynaptic motoneuron firing when receiving SCS-elicited sensory inputs alone. As shown in our simulations (**Extended Data Fig. 3b**), if SCS produces synchronous volleys in the recruited afferents, motoneurons should produce spikes aligned to these sensory inputs. We delivered SCS at a high frequency (100 Hz) in both monkeys in the same anesthetized procedure, thereby isolating motoneuron firing from alternative sources of modulation other than SCS. We then identified intraspinal single motor units from the ventral channels in the linear probe (**Fig. 5g**), corresponding to the location of motoneuron pools in lamina IX^34,35^. We computed ISI distributions and detected spectral peaks at exact integers of SCS pulses intervals (**Fig. 5h**). In both monkeys, motoneuron action potentials occurred near SCS pulses.

Third, we sought to confirm the link between SCS parameters and motoneuron firing modulation. Simulations revealed that the number of skipped SCS pulses increased with the SCS frequency (**Extended Data Fig. 3c-g**). Therefore, we computed the ISI cumulative distributions, which reflected this pattern (**Fig. 5j**). Indeed, for low SCS frequencies (30 Hz), sorted motor units skipped less SCS pulses in both monkeys and, in turn, the cumulative distribution of the ISI increased faster for less stimulation pulses in comparison to 100 Hz. We expanded this analysis for medium frequencies (60 Hz) and found consistent results in Mk-Ka and less pronounced in Mk-JC. Lastly, we computed the overall firing rate for the pool of sorted motor units and found that maximum firing activity was obtained at higher stimulation frequencies (i.e, 100 Hz in Mk-Ka and 60 Hz in Mk-JC, **Fig. 5i**). Hence, as predicted by the model (**Fig. 2d**), despite pulse-skipping at these high stimulation frequencies, motoneurons firing increased.

These experiments validated some of the model assumptions and results. However, being the monkeys anesthetized, we could not demonstrate our central results concerning combined volitional input and SCS and, specifically, the transformation of subthreshold SCS-induced EPSP into suprathreshold events by the residual supraspinal inputs.

### Volitional control of spinal motoneurons in humans during SCS

To verify our main findings concerning concurrent natural activity from supraspinal fibers and SCS, we conducted experiments in humans that received spinal cord implants. Specifically, we leveraged our existing Institutional Review Board protocols in two participants with poststroke arm hemiparesis (SCS03, SCS04), who participated in a pilot clinical trial exploring the effects of cervical SCS on motor control of the hand and arm^6^ **(Extended Data Fig. 6a,b)**, and an observational study in a participant with severe spinal cord injury (STIM01, ASIA A), who received an SCS implant on the lumbosacral spinal cord to reduce neuropathic pain in the lower limbs (**Extended Data Fig. 6c,d**). By testing our hypothesis in humans with stroke and spinal cord injury (SCI), we aimed at demonstrating that there are fundamental principles underlying the mechanisms of SCS irrespective of the specific disease that caused the lesion to the descending pathways.

We started by testing the effects of subthreshold SCS on motoneuron firing rates in STIM01, who had complete leg paralysis in consequence of a SCI; hence, a severe lesion. We asked the participant to contract his paralyzed leg in an isometric task at the maximum voluntary strength with and without SCS. Concurrently, we extracted motor units using surface EMG^36^ (**Fig. 6a,b; Extended Data Fig. 7a**). While the participant could not produce any force without SCS, SCS led to substantial force production (**Fig. 6c; Extended Data Fig. 8a**). However, the stimulation amplitude required to enable this voluntary production was just within the suprathreshold region. Indeed, we found significant motor responses even in moments in which the leg was supposed to be at rest (**Extended Data Fig. 8a**, brown area). When we inspected the ISI distribution of motoneurons during the isometric contraction task, we found clear peaks aligned to SCS pulses (**Fig. 6d; Extended Data Fig. 8a**) that closely resembled our simulations (**Fig. 2e,f**), which could not be explained by stimulation artifacts (**Extended Data Fig. 7d).**

**Fig. 6.**
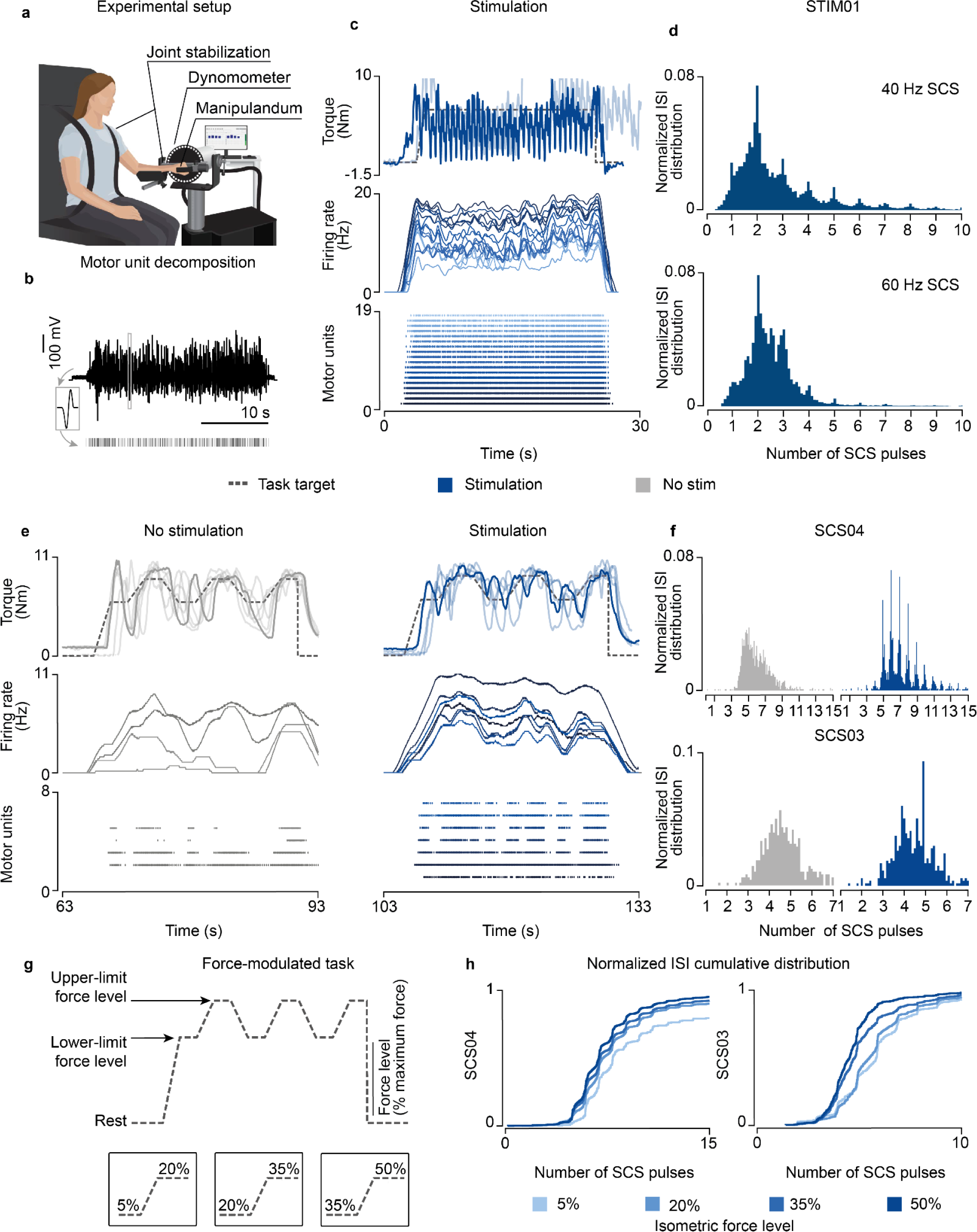
Experimental validation in humans with paralysis. **a,** Schematic of the experimental setup in participants with upper-limb paralysis performing a force task with their paretic arm (same setup was adapted for the participant with lower-limb paralysis). **b**, Schematic of motor unit decomposition using surface or high-density EMG performing a voluntary movement along with a sample raster plot for a detected motor unit waveform. **c**, Example of torque traces (top, overlapped with a trace from another repetition), firing rate (middle) and raster plot of the motor units (bottom) of both extensor muscles (Vastus Lateralis, Rectus Distal, sorted by average firing rate) during the isometric force task (35% of maximum force) in STIM01 during 40 Hz SCS. **d**, Histogram of ISI normalized probability distribution of the extracted motor units in STIM01 during 40 Hz SCS (above, 2 repetitions) and 60 Hz SCS (below, 2 repetitions). **e,** Example of torque traces (top, overlapped with trace from all repetitions), firing rate (middle) and raster plot of the motor units (bottom) during the force-modulated task (35-50% of the maximum force) in SCS04 with (60 Hz SCS) and without stimulation. **f**, Histograms of ISI normalized probability distribution of the extracted motor units in SCS04 (top, 18 repetitions) and SCS03 (bottom, 4 repetitions) with and without stimulation during 50% force-level period. **g,** Schematic of the parametrized isometric force-modulated task between pairs of force levels (force level is represented as percent of maximum force). Its lower and upper-limits were modified to perform a 5-20% maximum force task, 20-35% and 35-50%. **h**, Normalized ISI cumulative distribution of the extracted motor units during SCS in SCS04 (left) and SCS03 (right) during SCS for all force levels for all trials (18 repetitions for SCS04 for all for levels, 9 repetitions in SCS03 for 5% isometric force level, 7 for 20%, 7 for 35% and 4 for 50%).

We hypothesized that similar mechanisms should also apply for the control of upper-limbs, which are known to be more dependent on direct corticospinal input^6^. Thus, we tested our predictions in two participants with stroke (SCS03, SCS04) that received a cervical spinal cord implant. Our participants with stroke retained the ability to produce force also without SCS representing a milder lesion severity. We exploited this capability to test the validity of our model assumption that describes the firing of supraspinal fibers as stochastic (**Fig. 1b**) in natural conditions. As depicted by our simulations **(Extended Data Fig. 3a**), if supraspinal fibers fire stochastically, motoneuron firing should show non-synchronous firing patterns during volitional movement alone^37,38^ from the computed ISI distributions from single motor units from the paretic arm muscle (**Extended Data Fig. 7b,c,e**). Without SCS, as expected, the ISI distributions did not depict peaks of activity across time (plotted as a function of the number of SCS pulses for comparison purposes, **Fig. 6f**). Instead, during stimulation, once again the ISI distribution matched our findings with our simulations and in the SCI participant (**Fig. 6f; Extended Data Fig. 8b**). We could not test the involuntary SCS parameter regime as high stimulation amplitudes induced pain in these participants.

Lastly, we tested our model prediction that residual supraspinal inputs can modulate motoneuron firing rate and, thus, force by controlling the number of SCS pulses skipped. Thus, we sought to measure the motor unit firing for different force levels, which correspond to different levels of supraspinal input strength (**Fig. 6 e,g**). In particular, we parameterized the lower and upper force boundaries of our force-modulated task, which represent a percentage of the maximum force, and asked the participants to follow a target trace between 5-20%, 20-35% and 35-50% of their maximum force (**Fig. 6g**). The ISI cumulative distributions obtained from this task showed that both SCS03 and SCS04 skipped less SCS pulses when producing high forces, in contrast to low forces (**Fig. 6h; Extended Data Fig. 8c**).

In summary, our results in humans show that motor unit firing is aligned to stimulation pulses, regardless of motor diseases and lesion severity. We also demonstrated that, when present, residual supraspinal inputs can modulate motor unit firing induced by SCS to grade force production by reducing pulse-skipping.

### Subthreshold SCS efficacy may depend on lesion severity

In our human data we observed that, while stroke participants retained the ability to largely modulate SCS output, our SCI participant required high-stimulation amplitudes to enable movement that were already slightly suprathreshold. Therefore, we sought to exploit our biophysical model to study the theoretical impact that lesion severity could have on the range of effective parameters of SCS. We replicated in our model an isometric contraction task for a range of lesion severities (73-96%) and found that increasing the lesion severity to the supraspinal fibers reduced the combinations of subthreshold SCS parameters that generated sustained motoneuron firing (>8 Hz, **Fig. 7a**) with concurrent activation of the residual supraspinal inputs. In turn, this pattern was translated into the shrinking of the voluntary movement regime to smaller sizes (from 87 to 15 combinations of SCS parameters, **Fig. 7b,c; Extended Data Fig. 9a**).

**Fig. 7.**
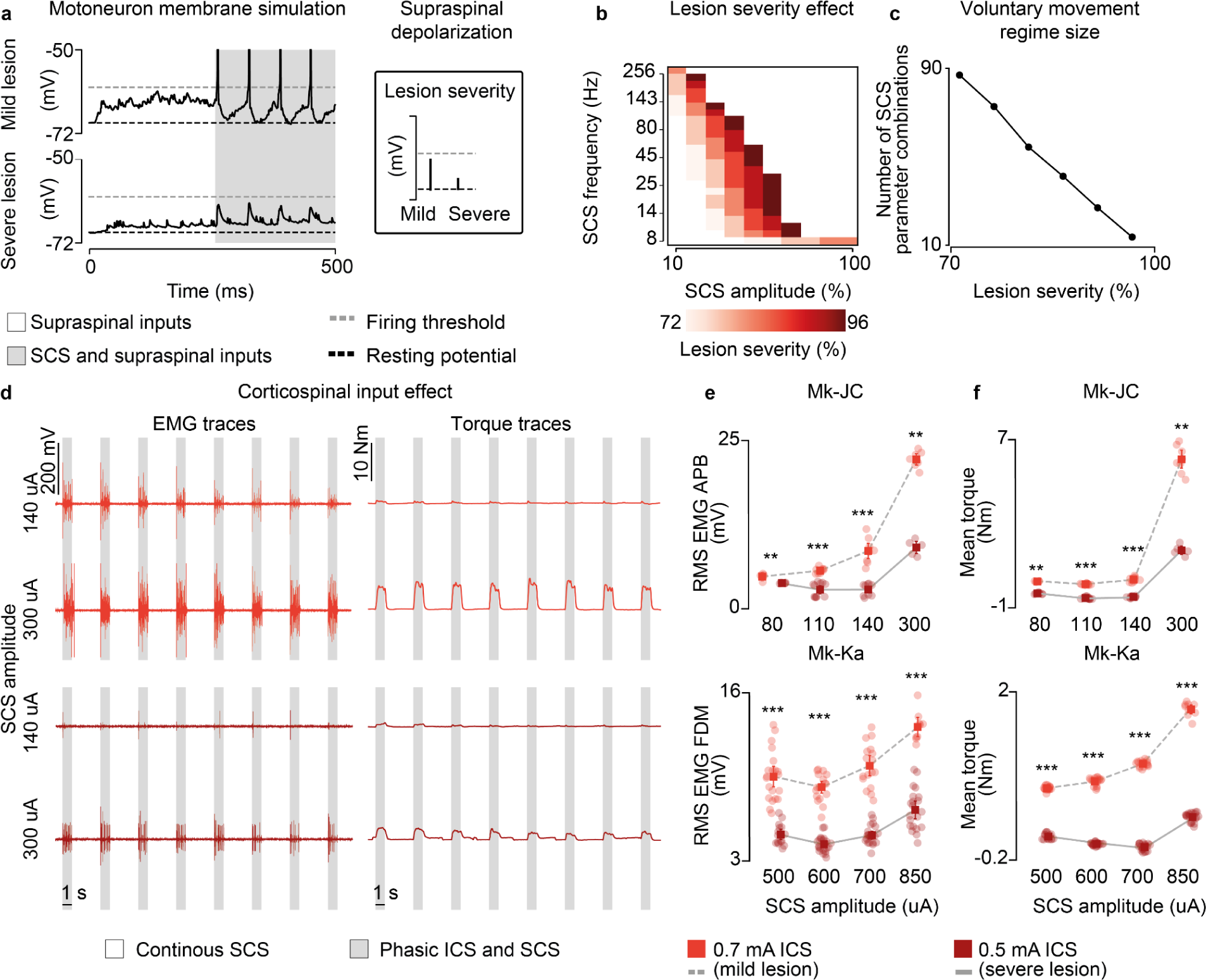
Lesion severity reduces voluntary movement regime. **a,** Simulation of the membrane potential of a single motoneuron driven by residual supraspinal fibers and SCS (16 Hz, 30%) in a mild (30 supraspinal fibers, top) and severe lesion (11 supraspinal fibers, bottom). **b**, Simulated heatmap indicating the combinations of SCS parameters within the voluntary movement regime for different degrees of lesion severity (mildest lesion in light red, the most severe lesion in dark red; 5 seconds simulation). **c**, Number of SCS parameter combinations within the voluntary movement regime for different degrees of lesion severity. **d**, Raw EMG and torque traces to illustrate the stimulation protocol employing phasic ICS (47 Hz) during continuous subthreshold SCS (60 Hz) at increasing SCS amplitudes (in Mk-JC, APB muscle). **e**, Root-mean square (RMS) of the EMG responses during phasic ICS and continuous SCS at increasing SCS amplitudes. Statistical quantification of the RMS EMG responses (***p<0.001; **p<0.01; Kruskal-Wallis test with 5, 5 points for 80 uA for 0.7 mA ICS and 0.5 mA ICS in Mk-JC; 10, 11 points for 110 uA; 9, 9 points for 140 uA; 7, 7 points for 300 uA; 21, 16 points for 500 uA for 0.7 mA ICS and 0.5 mA ICS in Mk-Ka; 18, 11 points for 600 uA; 17, 21 points for 700 uA; 9, 21 points for 850 uA). **f**, Mean torque during phasic ICS and continuous SCS at increasing SCS amplitudes. Statistical quantification of the mean torque (***p<0.001; **p<0.01; *p<0.05; Kruskal-Wallis test with 5, 9 points for 80 uA for 0.7 mA ICS and 0.5 mA ICS in Mk-JC; 10, 11 points for 110 uA; 9, 9 points for 140 uA; 8, 5 points for 300 uA; 18, 15 points for 500 uA for 0.7 mA ICS and 0.5 mA ICS in Mk-Ka; 18, 20 points for 600 uA; 16, 20 points for 700 uA; 9, 20 points for 850 uA). Square and error bars indicate the mean distribution of the data with 95% confidence interval.

We tested these results again in monkeys Mk-JC and Mk-Ka, where we could causally control the amount of inputs from the corticospinal tract via ICS. We superimposed subthreshold continuous SCS to bursts of ICS (**Fig. 7d**) and found that EMG responses produced during ICS bursts were potentiated as a function of the SCS amplitude. We then repeated the same stimulation protocol but at a very low ICS amplitude, thereby mimicking a case with very limited residual corticospinal inputs (i.e., a severe lesion). In this case, the root-mean square of the EMG responses decreased but it could be restored at higher SCS amplitudes, as in the case of our human patient with SCI (e.g., in Mk-JC, 8.4 mV during 140 uA SCS at 0.7 mA ICS vs 8.8 mV during 300 uA at 0.5 mA ICS, **Fig. 7e; Extended Data Fig. 10**), which was also clear in the mean torque produced by the hand grip (**Fig. 7d,f**).

Together, these results show that the useful parameter space at which SCS can facilitate production of volitional muscle activity shrinks with the reduction of the residual supraspinal inputs, thereby suggesting that the efficacy of SCS in facilitating motor control is inversely proportional to the severity of the damage to supraspinal pathways.

## DISCUSSION

In this study, we combined biophysical modeling with animal and human experiments to study the neural mechanisms underlying the recovery of voluntary control of spinal motoneurons during SCS after paralysis. We found that, after a lesion, residual supraspinal inputs can tonically depolarize spinal motoneurons bringing transient EPSPs induced by subthreshold SCS into action potentials. This transformation causes motoneuron firing to be aligned to SCS pulses, but supraspinal input can still influence motoneuron activity by controlling the number of active motoneurons and skipped SCS pulses. However, supraspinal control of motoneuron firing only occurs for a restricted range of subthreshold SCS parameters and the space of effective parameters shrinks for more severe supraspinal axon loss. Our results imply that the efficacy of SCS depends on the amount of spared supraspinal input.

### On the validity of the concept of neuromodulation

It is a common tendency to define the enabling effect that we describe here as “neuromodulation”. However, our findings exclude the possibility that SCS, delivered at frequencies below 250 Hz, is modulating the membrane of neurons in the strict sense of the term “modulation”, i.e., by biasing membrane excitability. In fact, it is the residual natural activity that can exert this biasing action by means of fast neurotransmitters such as glutamate, but also by the slower serotonergic and noradrenergic systems^39^. This natural biasing of cell excitability will then transform subthreshold, discrete SCS events into suprathreshold events (**Fig. 2**). While from behavioral observations, both interpretations are indistinguishable (since movement can only occur when intention is present), the specific mechanism does imply important differences on motoneurons behavior. For example, our model explains why stimulation frequency is related to proportional changes in strength and kinematics observed in our experiments (**Fig. 6**) and others^3,6,29,40,41^: SCS determines motoneuron spike times, forcing them to fire coherently with stimulation pulses. However, at the same time, residual supraspinal inputs can control the number of SCS pulses transformed into action potentials, thereby modulating motoneuron firing rates and producing controllable forces. This is in contrast to other neurotechnologies that directly drive motoneurons^42,43^, which would instead produce highly synchronized motoneuron firing and, in turn, result in poorly controllable forces. In other words, this is one of those cases in which the simple description of behavior does not offer satisfactory explanatory power of a phenomenon that requires analysis of neural output to be correctly interpreted^44^. In summary, while our results did confirm the “enabling” effects of SCS, they also suggest that the term “neuromodulation” should be used with caution, particularly, when studying the effects at the postsynaptic level, and suggest that similar effects may occur with other neurostimulation technologies like SCS for pain or deep-brain stimulation (DBS).

### Supraspinal control of movement after paralysis

Through the analysis of the membrane mechanisms of the response of spinal motoneurons to SCS pulses, we studied the computational principles that govern the integration of SCS mediated inputs to motoneurons and residual supraspinal activity. However, beyond spinal motoneurons, these mechanisms are likely to hold also for spinal interneurons that receive significant inputs from Ia afferents activated by SCS^45,46^. For example, central pattern generators^47,48^, inhibitory interneurons^49^ and excitatory premotor interneurons involved in muscle synergies^50^, all receive strong sensory inputs from proprioceptive afferents. Therefore, similarly to what we observed in motoneurons, residual supraspinal modulation can bring subthreshold SCS events into suprathreshold action potentials also in these network elements. In other words, the divergent connectivity of Ia afferents towards large parts of the sensorimotor network guarantees the brain access to many crucial components. This interpretation could explain why, despite the well-known low specificity of SCS^11,17^, we can observe repeatable, strong clinical effects^4,5^. In fact, we can speculate that a more selective technology that only enables control of motoneurons, or other specific sets of neurons, would fail at restoring motor control. The success of SCS does not stem from the activation of specific neural elements, but rather from the widespread occurrence of the membrane mechanism that we identified here in many spinal network nodes via the projection of activated sensory afferents, thus enabling complex, volitional motor control.

### Clinical implications

One of our most critical findings is that the well-defined SCS parameter regime that enables volitional control of spinal motoneurons shrinks as the number of residual supraspinal inputs decreases, i.e., as the severity of the lesion increases (**Fig. 7**). In other words, in a mild lesion (say 70% damage), where sufficient residual supraspinal fibers survived, the space of effective SCS parameters can be more than 7 times larger than in a severe lesion (say 95% damage). This is caused by the fact that, to compensate for severe loss of excitatory drive and to produce sufficient firing to enable movement, SCS amplitude must be set to values that rapidly enter the “suprathreshold” region, which we termed “involuntary movement” regime, meaning that activation and, likely, muscle co-contraction, will occur at rest. This intrinsic limitation of SCS emerges when inspecting aggregated experimental human data^51^ that show a clear trade-off between efficacy and lesion severity in SCI. In this scenario, the residual ability of people with paralysis to inhibit unwanted SCS inputs, or the possibility to acquire it with physical training, becomes more important than their capacity to excite spinal circuits (**Extended Data Fig. 9b)**. In fact, it is conceivable that a person with severe paralysis could learn to sculpt suprathreshold SCS inputs either by presynaptic inhibition of the Ia afferents or postsynaptic inhibition of the motoneurons, thereby suppressing unwanted activity and directing SCS excitatory drive towards movement-relevant neural pathways. It is possible that this is one of the effects of physical training, which, by strengthening synaptic connectivity, can enable voluntary control of SCS^2,8,18^. Finally, technology can also improve the efficacy of SCS, for example, closed-loop, spatiotemporal modulation of SCS^3,29,41,52^ can help support the sculpting of SCS and facilitate people with severe injuries to produce complex movements (**Extended Data Fig. 9c)**. While these alternatives can help increase the applicability of SCS, it is important to highlight that intrinsic limitations will eventually reduce SCS applicability and effectiveness for people with severe paralysis irrespective of the specific condition causing paralysis. It is conceivable that these limitations will be even more relevant for the recovery of upper limb movements given the strong dependence of hand control on corticospinal drive.

### Limitations

In this study, we only considered monosynaptic connections to spinal motoneurons coming from supraspinal axons as well as Ia afferents. Obviously, many more components that play a role in regulating excitability in the cord are stimulated by SCS, for example, large cutaneous mechanoreceptors, Ib afferents and group II afferents^10,11^. Nevertheless, as we discussed above, the basic membrane mechanism that regulates the changes in the membrane are likely to hold when considering these other inputs and, importantly, for other types of neurons that receive inputs from these afferents, such as Ia-inhibitory interneurons, central pattern generators and excitatory premotor interneurons implementing muscle synergies. Finally, our model only focuses on immediate effects of SCS and cannot be used to directly predict long-term changes that occur after physical training. Future work should be directed towards modeling long-term circuit changes that represent a major driver of SCS clinical efficacy.

In conclusion, we believe that detailed understanding of the neural mechanisms that drive the recovery of voluntary motor control will enable the optimization of SCS and other neurotechnologies to maximize efficacy of the residual brain-spinal architecture and control movement after paralysis, thereby promoting true motor recovery.

## Data Availability

All data produced in the present study are available upon reasonable request to the authors.

## ACKNOWLEDGMENTS

The authors would like to thank Prof. John Krakauer for his insightful inputs. The authors wish to thank Dr. Amr Mahrous and Dr. Vahagn Karapetyan for providing support during the monkey surgeries. The authors would also like to thank Isabella Bushko for the design of the monkey figure.

## AUTHOR CONTRIBUTIONS

JMB, GPO, MC conceived the study, designed the computational model and JMB performed the simulations. MC, DW and EP secured funding. PG, JGM, DF performed the surgery in monkeys and JMB, MC, LL, JH, EG, AD, EP performed the experiments. JMB, MC, LL, JH, EG, EP designed and implemented hardware and software for the monkey experiments. JMB, MC, LL, JH, PG designed the neurosurgical approach for the spinal implants. JH, LL, JGM, EP designed the neurosurgical approach for the brain implants. JMB, MC, LL collected all monkey data with the assistance of JH, EG, AD, EP. PG, DF performed the surgery in humans and JMB, GPO, MC, LF, SE, NV, ES, RdF, LB performed the experiments. GPO, MC, SE, SD, NV, ES, RdF, LB, PY designed and implemented hardware and software for the human experiments. MC, NV, ES, RdF, LB collected data in stroke participants and JMB, GPO, MC, LB, SE, SD in the SCI participant. NV, PY, DW performed motor unit decomposition collected from data in stroke participants. JMB, GPO performed the data analyses and prepared the figures. JMB, GPO, MC wrote the manuscript and all the authors contributed to its editing. MC supervised the work.

## COMPETING INTERESTS

MC and DW hold patents in relation to spinal cord stimulation. MC and DW are the founders of Reach Neuro, a company developing spinal cord stimulation technologies for stroke. EP has interest in Reach Neuro because of marital status with MC. All other authors declare they have no competing interests.

## FUNDING

This study was supported by start-up funds to MC from the Department of Neurosurgery, University of Pittsburgh, start-up funds to EP from the Department of Physical Medicine and Rehabilitation, University of Pittsburgh, and NIH grant number UG3NS123135 to MC and DW.

## METHODS

### Biophysical modeling

We implemented a biophysical model of motoneurons, Ia afferents and supraspinal fibers in Python 3.8 using NEURON 7.8^53^. The following number of neurons and their properties have been validated in similar models^10,11,24,25,27^.

#### Motoneuron model

We simulated a pool of 169 motoneurons. Each motoneuron comprised an integrated multicompartmental soma connected to an electronic-equivalent dendritic tree and myelinated axon, through an initial axon segment The established approach to describe the dynamics of the motoneuron membrane potential as a function of the ion channels is the Hodgkin-Huxley model. Equation 1 describes the membrane potential (V):

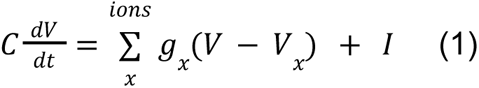

where C is the membrane capacity, *I* is current artificially injected to the neuron and *g_x_* is the conductance of the ion *x*, which depends on the membrane potential and ion concentrations. Thus, we implemented dedicated Hodgkin-Huxley equations for multiple ion channels (L-type Ca2+, Ca2+-activated K, delayed rectifier K+, N-type Ca2+, and nonlinear fast Na+) to study the integration of postsynaptic potentials from multiple presynaptic sources of modulation in the motoneuron membrane. Provided the increase in excitability in the motoneuron membrane after a lesion, we defined a model of postlesion motoneuron with modified ionic properties. Specifically, we reduced to 60% the Ca2+-activated K ion channel conductance^54^. We assumed that the motor cortex is unaltered after a lesion.

#### Excitatory synapses

Each motoneuron received excitatory inputs stochastically distributed along the dendritic tree from 60 Ia afferents, which were synchronously activated by each SCS pulse, and supraspinal fibers. The number of supraspinal fibers was defined by their synaptic weight and the number of pulses per second to each motoneuron^55^. In contrast to motoneurons, both EPSPs were modeled as point processes characterized by their conduction velocity and muscle fiber length. These synapses followed an exponential function with a reversal potential of *E_syn_* = 0.5 mV and a decay time constant τ = 0.5 ms.

Both EPSPs represented fast, glutamatergic synapses with similar dynamics to drive motoneuron excitation. Their synaptic weight followed a normal distribution (mean 0.047, standard deviation 0.0094)^24^. When supraspinal fibers point processes were activated, an EPSP was delivered in the motoneuron dendritic tree. To account for the variability in supraspinal fiber diameters, synaptic transmission delays followed a normal distribution (mean 0.25 ms, standard deviation 0.05 ms) plus a period of 1 ms. Instead, Ia afferents travel times were modeled as a latency between SCS pulses and elicited EPSPs that followed a log-normally distributed stochastic jitter (mean 1.0 ms, standard deviation 0.5 ms)^11^, thereby considering the difference in afferent diameter. Ia-afferent recruitment occurred when SCS pulses arrived outside the membrane refractory period. EPSP activation dynamics from both sources of modulation was modelled equally and followed a normal distribution (mean 1.6 ms, standard deviation 0.16 ms).

#### Force model

To estimate the force from the motoneuron pool firing rate, we used the model proposed in^30^. Here, we described the main equations (Equations 2-5). The twitch force generated for a spike from neuron *i* at time *t* is modeled as:

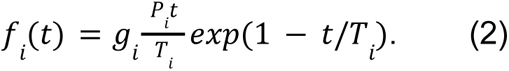

The peak twitch is defined as *P_i_* = *exp*(*b* · *i*) and the contraction time is *T_i_* = 90(1/*P_i_*)^1/4.2^. Finally, the gain, *g_i_* was modeled depending on the normalized mean interspike interval of neuron *i* (*ISI_i_*) and the contraction time *T_i_*:

For *Ti*/*ISIi* < 0. 4:

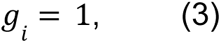

otherwise:

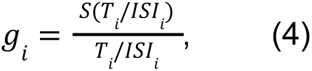

where *S*(*x*) is the sigmoidal function *S*(*x*) = 1 − *exp*(2*x*^3^).

Finally, the total normalized force is:

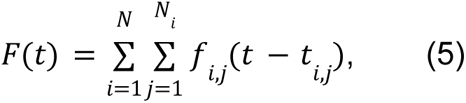

where *N* is the number of motoneurons and *N*_*i*_ is the number of spikes in neuron *i*. We always reported the force normalized with the maximum prelesion force.

### Experimental procedures in monkeys

#### Animals

All procedures were approved by the University of Pittsburgh Animal Research Protections and Institutional Animal Care and Use Committee (ISOOO17081). We conducted experiments in 2 anesthetized Macaca Fascicularis (Mk-JC: age 6, 7.5 kg, male; Mk-Ka: age 5, 6.8 kg, female). The animals were housed in the primate facility at the Division of laboratory Animal Resources at the University of Pittsburgh. The animals had unrestricted access to water and food as well as daily enrichments.

#### Surgical procedures

Animals underwent two surgical procedures. First, in a survival procedure, we acquired in vivo structural magnetic-resonance imaging and computed tomography for each animal. Collected imaging data was used to plan the trajectories of the deep-brain probe that stimulated the posterior limb of the internal capsule, which innervates the hand muscles. We further describe this procedure in^56^. Second, in a terminal procedure, certified neurosurgeons performed muscle implantation, robotic deep probe implantation and spinal probe implantation (described below in detail) while the monkeys had continuous ventilation support and were under full anesthesia induced with ketamine (10 mg/kg, i.m.) and under continuous intravenous infusion of propofol (1.8-5.4 ml/kg/h) and fentanyl (0.2-1.7 ml/kg/h). Anesthesia conditions were maintained until the conclusion of the experiments, where animals were euthanized with a single injection of pentobarbital (86 mg/kg, i.v.).

We inserted a pair of EMG needle electrodes (disposable single 7mm Subdermal needle electrode, Rhythmlink) into the hand muscles (abductor pollicis brevis (APB), flexor digiti minimi (FDM)). The reference was inserted in the lower back or the tail with a needle electrode. Subsequently, we fixed the monkey’s head in a stereotactic frame (Kopf, Model 1530, Tujunga, CA, USA) with the cervical spine fixed in a prone position.

We proceeded with the robotic deep probe implantation (ROSA robot)^56^. To accurately implant a probe in the hand area of the internal capsule (AC-PC level), a frontal insertion trajectory was guided by the robot software (ROSA One(R) Robot Assistance Platform). Trajectory planning was achieved due to previously collected imaging data^56^. The probe positioning was contralateral to the muscle implants. We precisely drilled a penetration hole in the skull for the implantation of a fixation bolt. Next we inserted a radiofrequency electrode (RF-SE-10 Reusable Stainless Steel Electrode, Abbott). We confirmed the correct position of the probe by inspecting the hand muscles EMG activity in response to ICS (2 Hz, 0.8-2 mA) that should produce monosynaptic activation of the cervical motoneurons.

Lastly, we performed a laminectomy from C3 to T1 vertebrae and exposed the cervical spinal cord to the nerves that innervated targeted muscles. After opening the dura matter, we placed a stimulator (bipolar, 73602-190/10, Ambu Neuroline, Denmark) on the cervical dorsal roots (Mk-JC: C8, Mk-Ka: C6). We confirmed its positioning by verifying EMG responses in the target muscles. Furthermore, we implanted a 32-channel linear probe (A1×32-15mm-100-177-CM32 Linear Probe with 32 pin Omnetics Connector, NeuroNexus, Ann Arbor, MI, USA) at the cervical spinal segment rostral to the stimulation site (Mk-JC: C7, Mk-Ka: C7) using Kopf micromanipulators (Kopf, Model 1760, Tujunga, CA, USA). To do so, we pierced the pia using a surgical needle to ensure safety probe penetration. Its placement was ∼1.2 mm parallel to midline (ipsilateral to the muscle implants) due to the flow of sensory information of Ia afferent in the dorsoventral direction consistent with the placement of the probe. Its length (3.1 mm) is designed to cover the gray matter of the monkey’s spinal cord and record extracellular potentials as well as intraspinal spiking activity. The reference was inserted under the skin next to the probe insertion site.

#### Grasp force data acquisition

We placed a force transducer in the anesthetized monkey’s hand (ipsilateral to the muscle implants). Grasp force data was captured with a 6-axis low-profile force and torque (F/T) sensor (Mini40, ATI Industrial Automation, North Carolina). The sensor was powered using a multi-axis (F/T) transducer system (ATI DAQ F/T) that also calibrated the force data. The sensor measured three degrees of force (+/-810 N for Fxy, +/-2400 N Fz) and torque (+/- 19Nm Txy, +/- 20Nm Tz) simultaneously. A small rod was mounted to the xy-plane of the sensor to be a grip handle for the animal.

#### Data acquisition and electrophysiology

We employed an AM stimulator (model 2100 A-M Systems, Sequim, WA, USA) for the stimulation of the dorsal roots and internal capsule as biphasic symmetric square pulses in the form of single or train of pulses and detailed pulse widths (Mk-JC: 0.40 ms for SCS, 1 ms for ICS; Mk-Ka: 0.45 ms for SCS, 0.8 ms for ICS). Specific stimulation configurations and parameters are defined in each figure. After data amplification (Nano2-HV headstage) and digital processing, neural recordings (EMG and intraspinal spiking data) were recorded (Ripple Neuro Grapevine Neural Interface 1.14.3) at 30 kHz sampling frequency along with the force/torque data (same sampling frequency). Through the data acquisition platform (Trellis), we developed an in-house interface that computed the EMG stimulation-trigger averaged responses in real-time to determine the stimulation amplitude for motor threshold responses for each muscle. Moreover, intraspinal recordings were treated with a broadband filter between 300 Hz and 3 kHz and a voltage threshold (3 root-mean square) to determine neural events.

#### Analysis of muscle activity

EMG signal was calculated for each muscle differentially between the two needle electrodes and was bandpass filtered between 30 and 800 Hz with a 2nd order Butterworth filter. We then identified the peak-to-peak amplitude of reflex-mediated responses evoked by the paired stimulation in a window of 25 ms after the SCS pulse. Moreover, we computed the root-mean square of the response during phasic ICS alone and concurrent phasic ICS and continuous SCS. This analysis was performed offline in MATLAB (version R2019b).

#### Analysis of single unit activity

We sorted spikes from the 8 most ventral channels of the 32-channel linear probe. Previously, we applied a comb filter of the raw signal to remove artifacts at 60 Hz. We then applied a 3rd order Butterworth digital filter (800-5000 Hz). Spikes were detected by waveform threshold crossing. All intraspinal recordings were concatenated by channel for comparison purposes across performed experiments. Using a spike sorting software (Plexon Offline Sorter, Dallas, Texas), we identified single motor units by channel with semi-automatic k-means clustering. Regarding the histograms of the normalized ISI probability distribution of the sorted units, we used a bin width of 0.1 ms. Previously, we noticed that the application of continuous SCS resulted in an initial short hyperexcitable period, thus, we discarded the first 5 seconds of spiking data after the start of the stimulation to perform our analysis on stationary conditions. This analysis was performed offline in MATLAB (version R2019b).

#### Analysis of torque activity

Mean torque signal was computed as the module of all three measured directions and filtered by applying a 15 kHz low bandpass filter with a 3rd order Butterworth filter. Provided that monkeys were anesthetized, we obtained the baseline value by subtracting the mean torque response during the no stimulation period during ICS alone for each amplitude (0.7 mA and 0.5 mA). This analysis was performed offline in MATLAB (version R2019b).

### Experimental procedures in humans

#### Trials information

All experimental protocols were approved by the University of Pittsburgh Institutional Review Board (IRB): STUDY19090210, for the stroke participants (SCS03 and SCS04), and STUDY22020031, for the spinal cord injury participant (STIM01). All participants provided informed consent according to the procedure approved by the IRB of the University of Pittsburgh and participants were compensated for each day of the trial and for travel and lodging during the study period.

The stroke participants (SCS03 and SCS04) were recruited to participate in an exploratory clinical trial to obtain preliminary evidence of safety and efficacy of SCS to improve motor control in people with chronic post-stroke upper-limb hemiparesis^6^. A detailed description of the trial, including inclusion criteria and study design, has been described in^6^ and can be found on <u>ClinicalTrials.gov</u> (<u>NCT04512690</u>). The spinal cord injury participant (STIM01) was recruited to participate in a basic research study enrolling people already living with an epidural spinal cord stimulator (STUDY22020031). During the experimental sessions in STIM01, we changed the stimulation parameters to perform the different tasks using the Medtronic Intellis Platform. At the end of each session, we reset the stimulation parameters to their clinical values.

#### Participants Information

STIM01 (Black male, 20-25 years old) was diagnosed as ASIA A after a spinal cord injury at T6 vertebra level 5 years ago. He was implanted with a spinal cord stimulation to reduce pain and spasticity 4 years ago. Since the implantation, he has combined SCS and physical therapy to regain control of his lower limbs. When he enrolled in the study, he was able to voluntarily move his left leg during SCS. STIM01 was implanted with a Medtronic paddle array from the bottom of T11 to the top L1 vertebrae. SCS03 (White male, 40-45 years old) suffered a hemorrhagic stroke followed by craniotomy 7 years before participating in the study and was diagnosed with a Fugl-Meyer motor score of 28. SCS04 (Black male, 40-45 years old) suffered an ischemic stroke in the anterior frontal and temporal lobe 5 years before participating in the study and was diagnosed with a Fugl-Meyer motor score of 19. To minimize risks in SCS03 and SCS04, participants were temporally implanted for 29 days, after which electrodes were explanted. IDs (i.e., STIM01, SCS03, SCS04) are not known to anyone outside the research group.

#### Torque experiments

We performed the torque experiments in all subjects using a robotic torque dynamometer (HUMAC NORM, CSMi) that can be set to measure torque in different joints. These experiments and data acquisition are described in detail in^6^.

For STIM01, we measured his maximum torque during knee extension for 6 repetitions of 2 seconds (**Extended Data Fig. 8a**). We also asked STIM01 to hold a torque level of 35% of his maximum torque during 30 seconds (**Fig. 6c**). In both tasks, we placed a 4-channel EMG sensor (Trigno Galileo Sensor, Delsys) to record EMG activity on the Vastus Lateralis and Rectus Femoris.

SCS03 performed an isometric force-modulated task for elbow flexion where he was asked to follow a predefined force trace ranging from 5-20%, 20-35%, and 35-50% of his maximum voluntary contraction force with and without SCS (**Fig. 6g; Extended Data Fig. 8c**). This experiment was repeated with and without SCS. A 64-channel high-density EMG grid (TMSi, The Netherlands) was placed over Bicep Brachii to record muscle activation during the experiment.

Because of SCS04’s inability to perform the force-modulated task at the elbow, he performed a similar task of grasp force-modulated task at different force levels, ranging from 5-20%, 20-35%, and 35-50% of his maximum torque, both with and without SCS (**Fig. 6g; Extended Data Fig. 8c**). The grasp force data was measured with a 6-axis low-profile force and torque (F/T) sensor (Mini40, ATI Industrial Automation, North Carolina). A 32-channel high-density EMG grid (TMSi,The Netherlands) was placed over the anterior forearm (specifically, Flexor Carpi Radialis) to record muscle activity during the task.

#### Motor unit decomposition

In STIM01, we used the Neuromap software (Delsys) to decompose the EMG activity in single motoneuron action potentials. For the experiments with SCS03 and SCS04, single motor unit firing rates were identified from high-density signals by the spike trains of motor units, which were identified by the Convolution Kernel Compensation (CKC) technique^57^. The high-density EMG signals were first bandpass filtered between 20 and 500 Hz with a 3rd order Butterworth filter. Additionally, SCS artifact was removed by notch filtering at the SCS frequency. SCS harmonics were also removed using notch filtering by identifying peaks in the power spectrum associated with SCS. The filtered high-density EMG signals were decomposed by the DEMUSE software (v6.1; The University of Maribor, Slovenia)^58,59^. The decomposed motor unit spike trains were processed to identify single motor units and calculate their firing rates. During this step, physiologically irregular motor unit firings (<5 Hz and >50 Hz) were discarded^60,61^. The irregular motor unit firings were identified by following the guidelines in literature and accuracy of identified motor units was determined by the Pulse-to-Noise Ratio (>25 dB)^62,63^ **(Extended Data Fig. 7b,c,e).**

#### Analysis of torque activity

To account for the weight of the paretic limb in STIM01, torque responses were baseline corrected with the mean resting torque for the first 60 ms. Moreover, given the fluctuations in the responses, we smoothed the data applying a moving window of 100 ms. Lastly, we computed the maximum torque during the maximum voluntary contraction periods for each repetition. Only for SCS04, given that the force-modulated task was performed with the same data acquisition system as in the monkeys, torque responses were analogously treated by applying the same low bandpass filter and baseline correction with the mean resting torque for the first 3.33 seconds. This analysis was performed offline in MATLAB (version R2019b).

#### Analysis of firing rate activity

Only for SCS03 and SCS04, we computed the firing rate in a 50 ms window with a sliding window of 5 ms over the entire duration of the trial and zeropadded for 45 ms and smoothed it by applying a moving window of 100 ms. We identified firing rates for each unit during a constant force level, i.e., a constant supraspinal innervation (replicating what we did in our biophysical model). Like in the monkey normalized ISI probability distributions, we used a bin width of 0.1 ms. As indicated in the captions, data included in the figures correspond to all the repetitions under the same conditions and session. This analysis was performed offline in MATLAB (version R2019b).

### Statistical analyses

All statistical comparisons were performed using the Kruskal-Wallis test, which is a nonparametric approach to the one-way ANOVA. Subsequently, followup multiple comparison tests on pairs of sample medians were performed. Data points outside 1.5 times the interquartile range plus the 25th/75th percentile were classified as outliers and discarded. For illustration purposes, the mean distribution of the data with 95% confidence interval was indicated for each distribution. This analysis was performed offline in MATLAB (version R2019b).

### Code Availability

Software routines developed for data analysis are available from the corresponding author upon reasonable request.

## DATA AVAILABILITY

The main data supporting the results in this study are available within the paper. All data generated in this study will be uploaded in a public repository upon acceptance of the manuscript. Raw data will be available upon reasonable request to the corresponding author.

## ADDITIONAL INFORMATION

**Extended Data Fig. 1.**
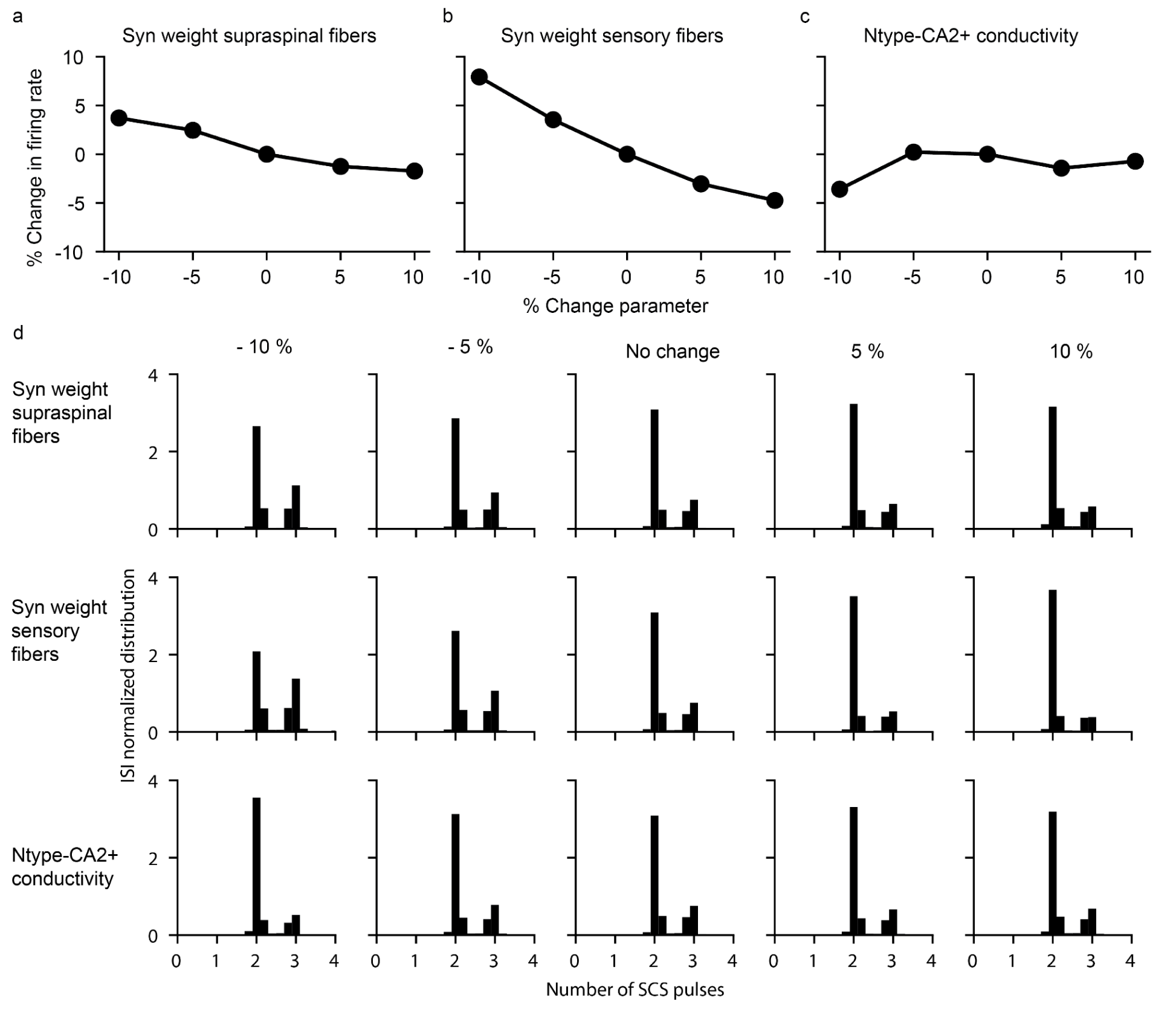
Motoneuron firing is robust to parameter changes after a lesion. **a-c,** Changes up to ±10% in key parameters of the model (mean synaptic weight from residual supraspinal fibers to motoneurons (**a**), mean synaptic weight from sensory fibers to motoneurons (**b**) and the conductivity of the N-type Ca2+ ion channel (**c**)) do not produce dramatic changes in motoneuron firing rate, thereby indicating that the model is robust to changes in key parameters. **d,** Normalized ISI probability distribution for the different modifications of the key parameters in **a-c**. The transformation of subthreshold SCS-pulses in suprathreshold events do not depend on the fine tuning of these parameters.

**Extended Data Fig. 2.**
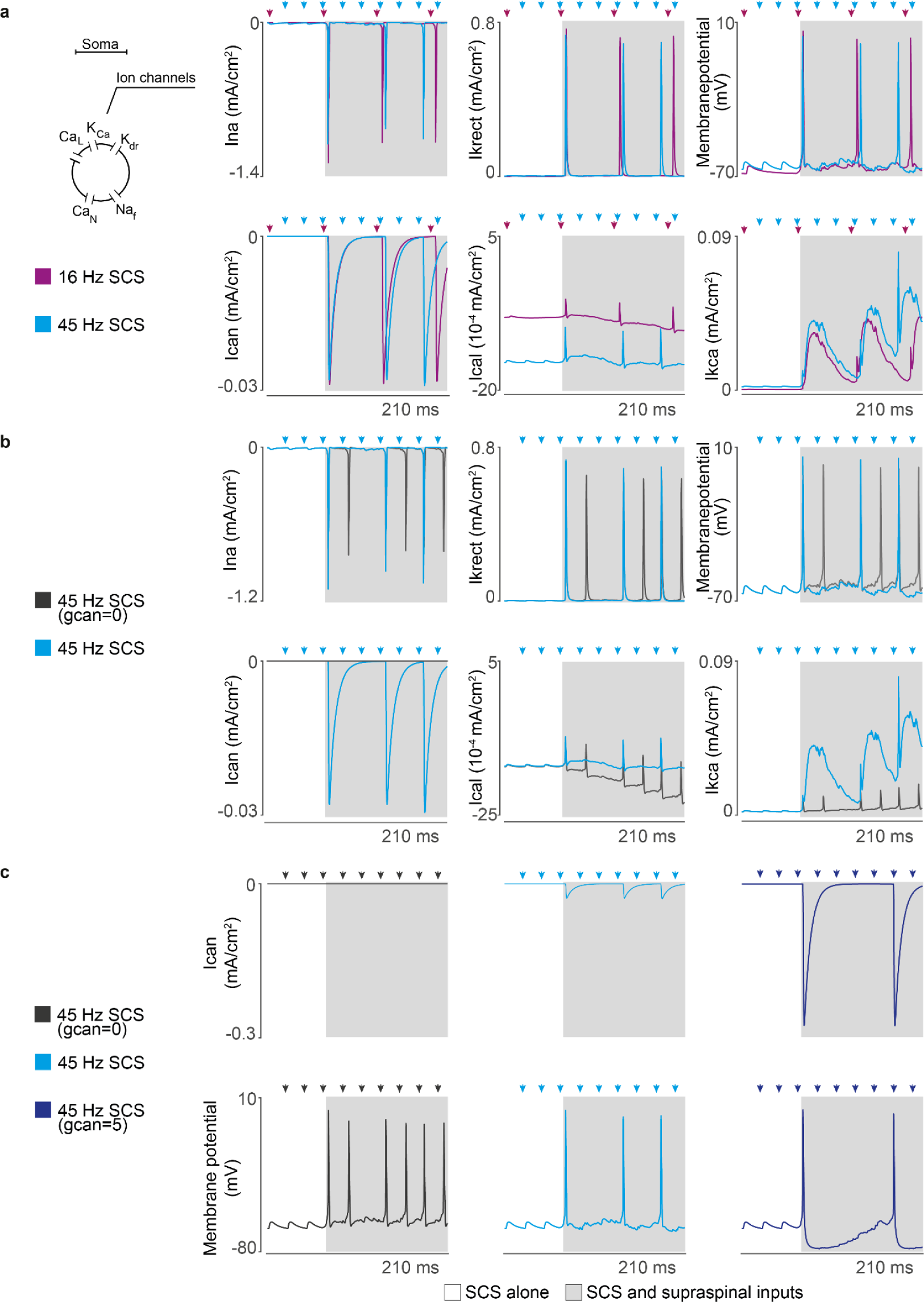
Motoneuron ion channel dynamics in our biophysical model. **a,** Ion currents of a single motoneuron in response to different SCS frequencies (same amplitude, 50%) along with the motoneuron membrane potential during concurrent innervation of residual supraspinal inputs (gray). Ionic currents: L-type Ca2+ (Ical), Ca2+-activated K (Ikca), delayed rectifier K+ (Ikrect), N-type Ca2+ (Ican), and nonlinear fast Na+ (Ina). At low stimulation frequencies (<19 Hz), motoneurons generate action potentials with concurrent supraspinal input for each stimulation pulse. At higher frequencies, motoneurons skip some stimulation pulses due to N-type Ca2+ slow time dynamics. **b**, Ion currents of the same motoneuron at 45 Hz SCS (50% amplitude) for different conductances of the N-type Ca2+ ion channel. Setting the conductance close to 0 relaxes N-type Ca2+ dynamics constraints. Consequently, less rectifier K+ ions leave the membrane to stabilize the motoneuron membrane potential, leading to a shorter refractory period in the overall motoneuron membrane and producing more action potentials following each stimulation pulse. **c**, Ion current of the N-type Ca2+ ion channel varying its conductance and its resulting membrane potential. When membrane dynamics allow the entrance of N-type Ca2+ ions, a cascade of Ca2+-activated K ions respond proportionally to counteract this entrance, leading to exaggerated refractory periods in the motoneuron membrane. Arrows indicate stimulation pulses.

**Extended Data Fig. 3.**
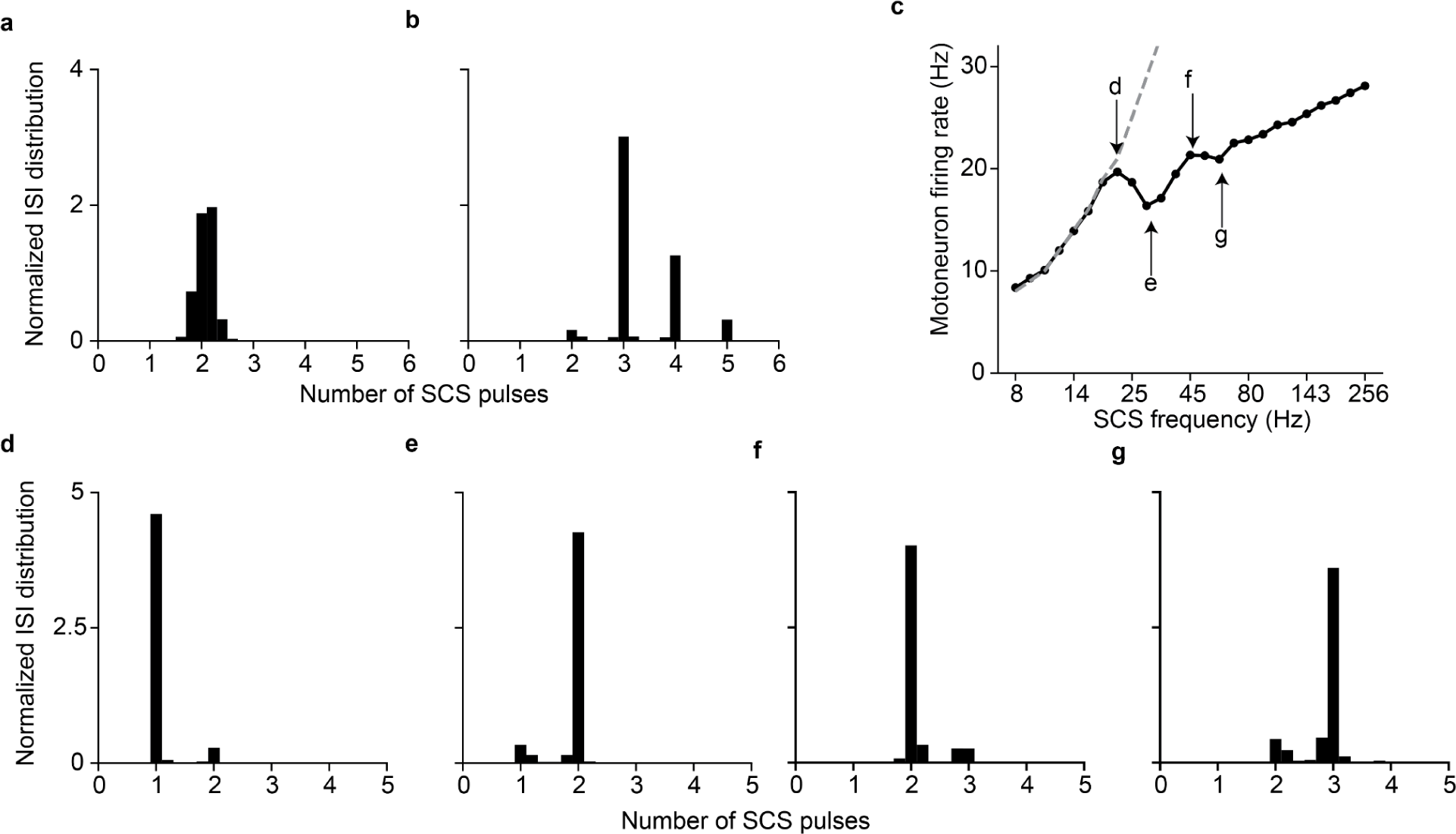
Motoneuron firing rate increases non-monotonically with SCS frequency. **a,** Normalized ISI probability distribution of a motoneuron pool prior to the lesion (i.e., without SCS) showing no peaks at integer numbers. For comparison purposes with panel **b**, the time is set in SCS pulses units with a stimulation frequency of 45 Hz. **b,** Normalized ISI probability distribution of the same motoneuron pool during SCS alone (45 Hz, 50% amplitude). Spikes reliably follow SCS pulses. **c,** Firing rates of a pool of motoneurons as a function of SCS frequencies (constant 60% amplitude). Motoneurons are concurrently receiving excitatory inputs from residual supraspinal fibers. **d-g,** Normalized ISI probability distribution of a motoneuron pool in SCS pulses units for different SCS frequencies: 19 (**d**), 29 (**e**), 45 (**f**) and 60 (**g**) Hz (note arrows in **c**). The different peaks in the Normalized ISI probability distribution is a consequence of transforming SCS pulses into action potentials in the motoneuron membrane. In general, motoneuron firing rate increases with SCS frequency (**c**). However, when motoneurons are unable to produce an action potential for each SCS pulse, the firing rate drops (see arrows **e** and **g** in panel **c**). Given the membrane dynamics, following an action potential, there is a refractory period that sets the ISI greater than the interpulse interval (IPI) and motoneurons skip SCS pulses. This skipping effectively decreases the firing rate of the motoneuron pool. Hence, in the local maximum marked with the **d** arrow, the ISI is the same as the IPI (**d**) (i.e., all SCS pulses trigger an action potential) while the minimum coincides where almost all motoneurons skip one pulse (**e**). However, the IPI decreases with higher SCS frequencies, producing an increase in firing rate between point (**e**) and (**g**). There is another local maximum in firing rate when motoneurons skip two SCS pulses (**f**), while again the minimum coincides where most motoneurons skip two pulses (**g**). Finally, at increasing SCS frequencies, the firing rate of the pool increases consequently.

**Extended Data Fig. 4.**
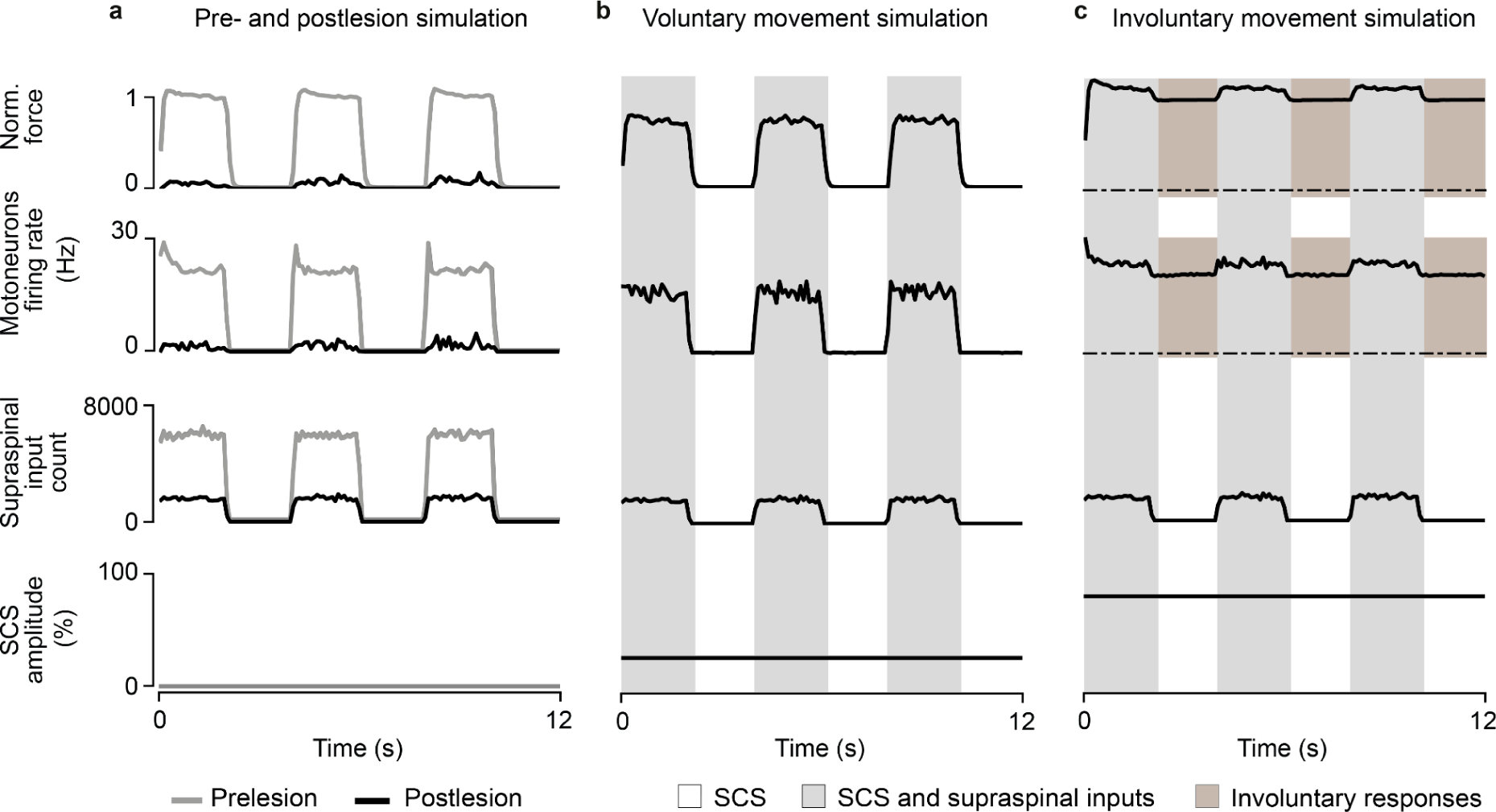
Simulations of SCS parameter regimes in an isometric task. **a**, Simulation of a pool of motoneuron pool during a maximum voluntary contraction task during phasic activation of supraspinal fibers (2 seconds active, 2 seconds inactive). Traces represent the normalized force (top), motoneuron pool firing rate (middle), supraspinal input count (bottom) in a pre- (gray) and postlesion (black) system. **b**, Same as **c** during continuous SCS at parameters belonging to the voluntary movement regime (SCS frequency 60 Hz, amplitude 30%). Motoneurons fire only coinciding with supraspinal activation, achieving a firing rate only 19% smaller than the prelesion case. Bottom trace indicates the continuous SCS amplitude. **c**, Same as **b** during continuous SCS at parameters belonging to the involuntary movement regime (SCS frequency 60 Hz, amplitude 80%). Motoneurons fire also when supraspinal fibers are silent.

**Extended Data Fig. 5.**
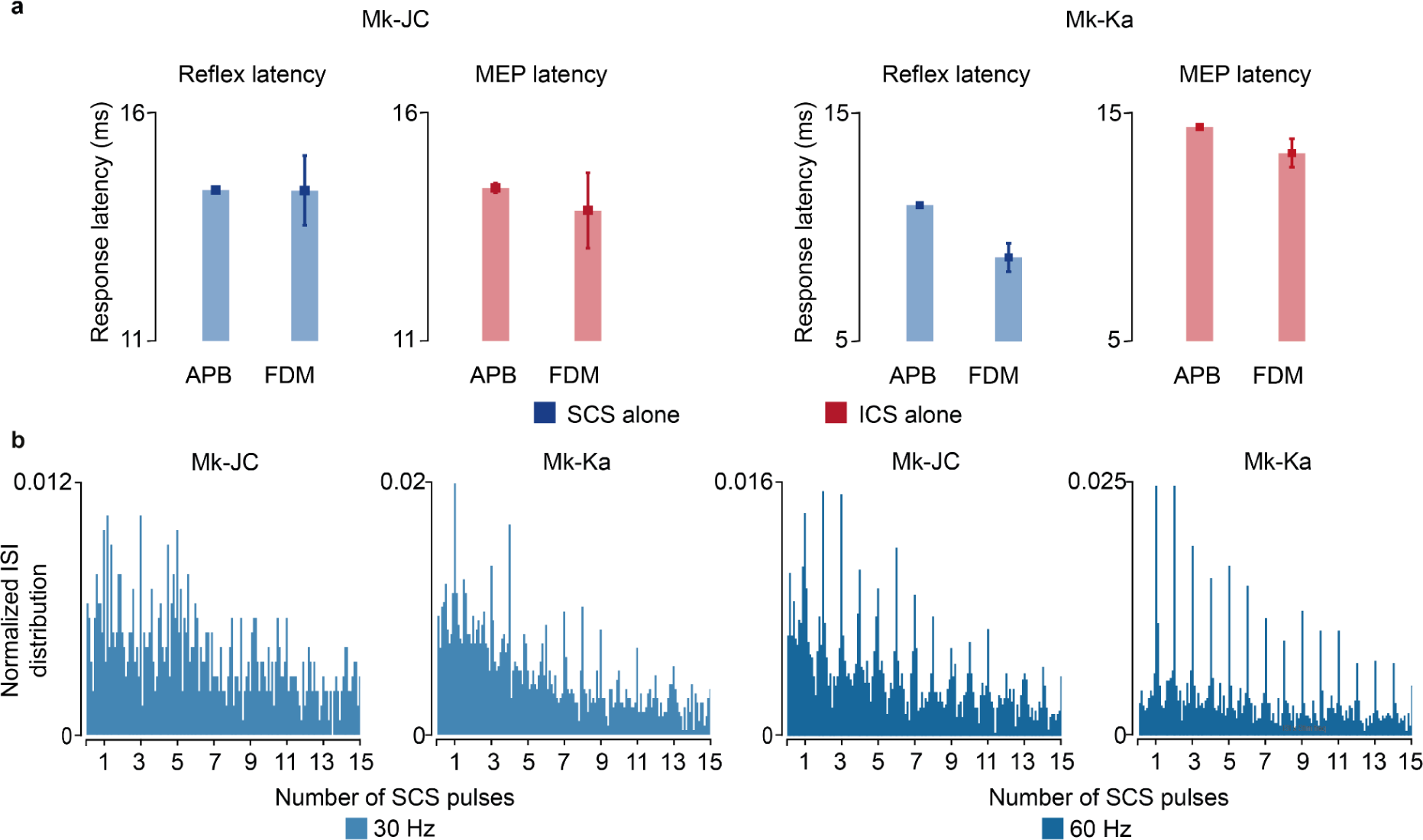
Monkey EMG response latencies and intraspinal ISI distributions. **a,** EMG response latencies for each monkey evoked by a single SCS pulse (H-reflex, blue) and ICS pulse (motor-evoked potential (MEP), red). **b**, Histograms of ISI normalized probability distribution of the sorted motor units during 30 and 60 Hz SCS in both monkeys. Square and error bars indicate the mean distribution of the data with 95% confidence interval.

**Extended Data Fig. 6.**
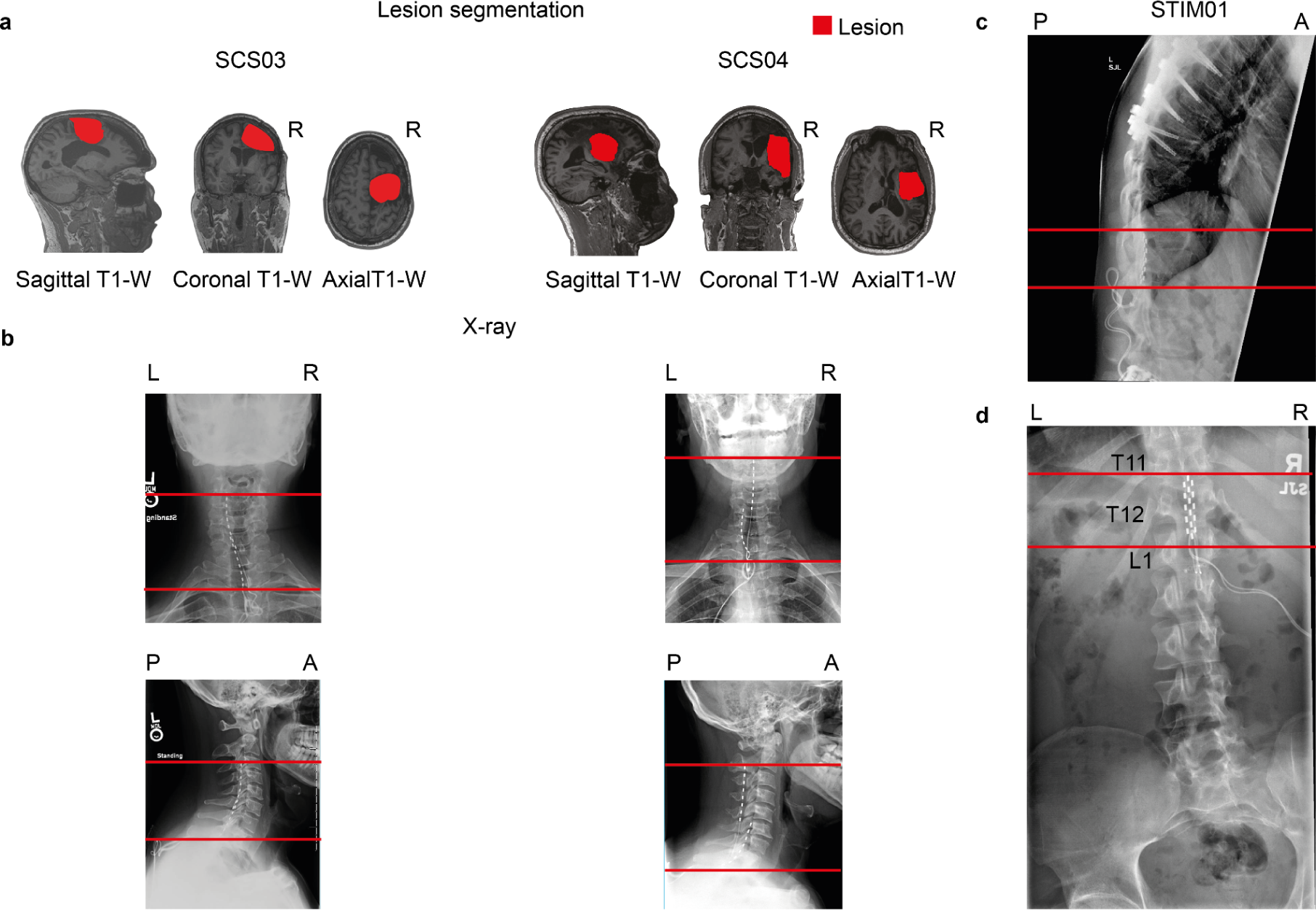
Lesion characterization and SCS implantation in humans with paralysis. **a,** Sagittal, coronal, and axial T1-weighted magnetic-resonance imaging (MRI) 2D projections for SCS03 and SCS04 using a 3T Prisma system (Siemens) using a 64-channel head and neck coil (T1-weighted structural image was captured using a magnetization-prepared rapid gradient echo sequence: repetition time = 2,300 ms; echo time = 2.9 ms; field of view = 256 × 256 mm2; 192 slices, slice thickness = 1.0 mm, in-plane resolution = 1.0 × 1.0 mm). The segmented lesion is shown in red for both participants and performed as described in^6^. R indicates the Right hemisphere. **b**, X-rays for SCS03 and SCS04 showing the position of the spinal leads. The red lines mark the same anatomical location across the X-rays to facilitate interpretation. Minimal displacement occurred after initial implantation. **c-d**, X-rays for STIM01 showing the position of the spinal leads and the lesion. The red lines mark the same anatomical location across the X-rays to facilitate interpretation. Minimal displacement occurred after initial implantation.

**Extended Data Fig. 7.**
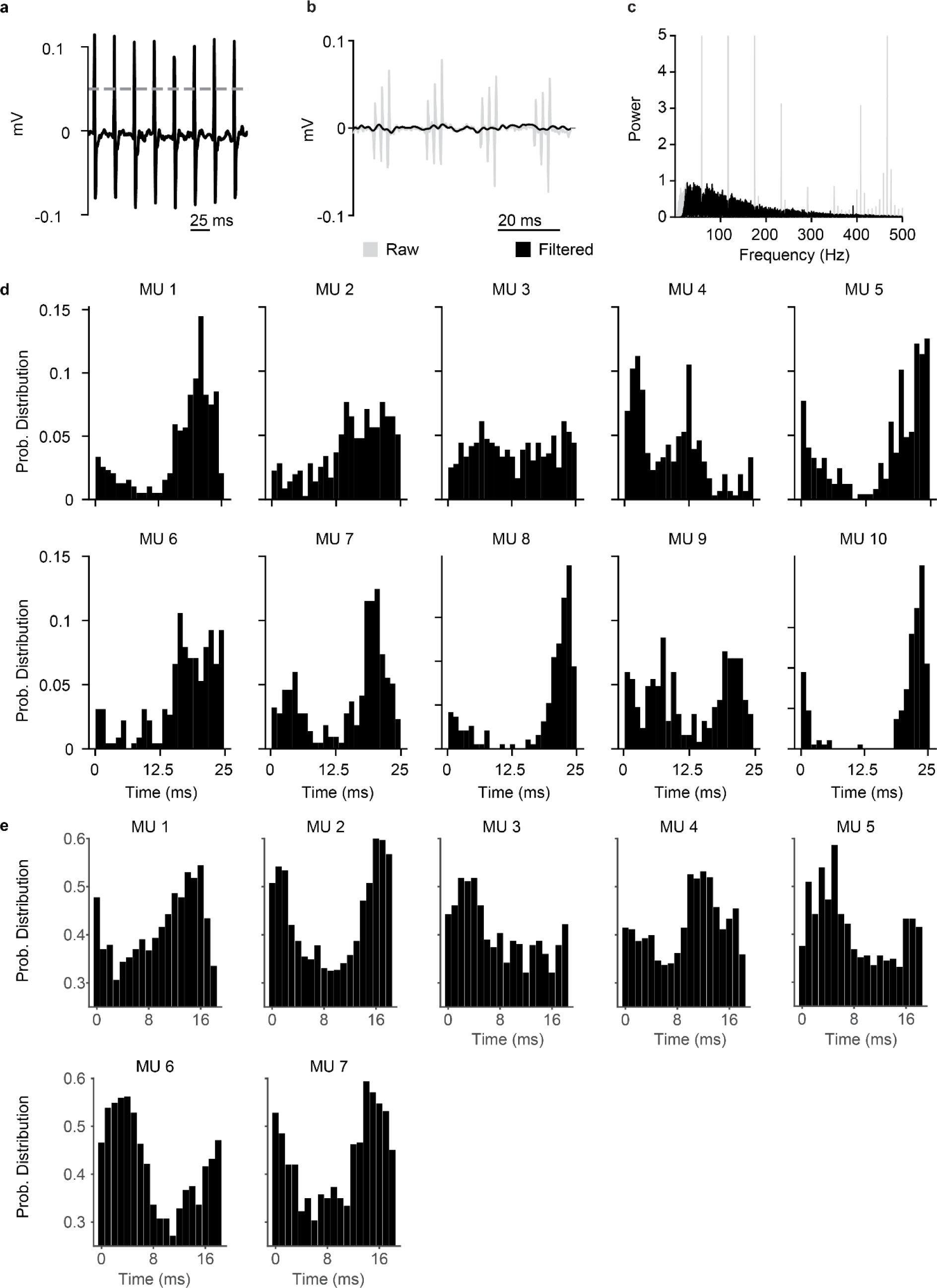
Action potentials triggered by SCS pulses are not stimulation artifacts in humans with paralysis. **a,** For the experiments with STIM01, we recorded the stimulation artifact with a surface EMG placed on the dorsal. We defined the stimulation timestamps through EMG threshold-crossing above 0.05 mV (dashed line). **b**, For the experiments with the stroke participants, traces were recorded from high-density EMG: in gray showing the presence of stimulation artifacts and in black after removing the stimulation artifacts (showing an example of SCS03, 60 Hz SCS). **c**, For the experiments with the stroke participants, stimulation artifact removal was achieved by applying a series of notch filters at harmonics of SCS frequency to remove peaks in the Fast Fourier transform (shown in gray) of the bandpass filtered signal (see methods). **d,** Examples of the probability distribution of the time between the most recent SCS pulse and an action potential for each motor unit (MU; 40 Hz SCS, 1 ms bin) using surface EMG in STIM01. If the action potentials detected by the acquisition software were stimulation artifacts, they would be aligned to the SCS pulse, thereby showing histograms with a high peak at time 0. Given that the times from the most recent SCS pulse and the action potentials are distributed with a mean between 12.5 and 25 ms, the action potential detected can not be artifacts of SCS. **e**, Same as **d** using the high-density EMG in the stroke participants (SCS03, SCS04; 60 Hz SCS, 1 ms bin).

**Extended Data Fig. 8.**
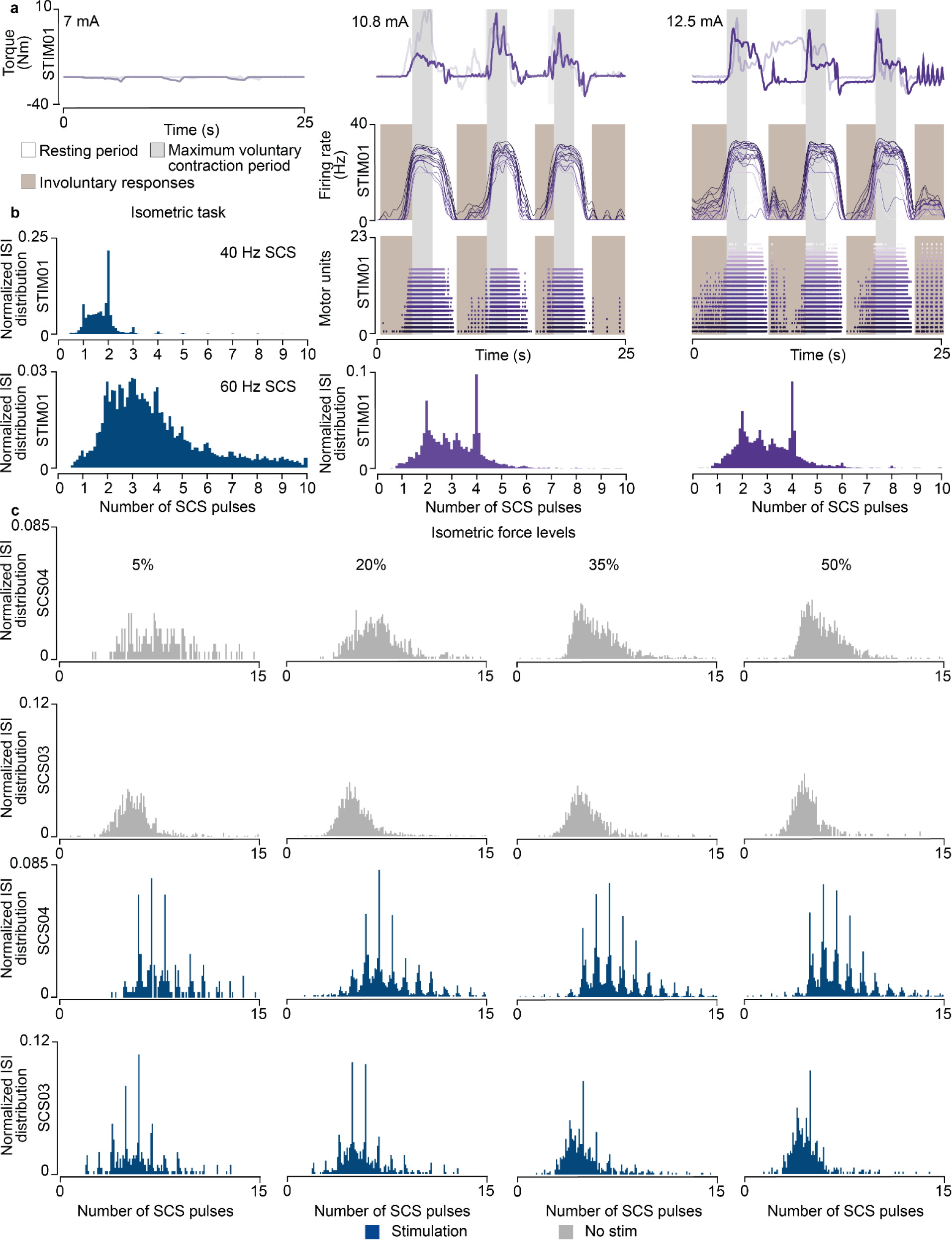
Experimental validation in humans with spinal cord injury and stroke. **a,** Torque traces (top, overlapped with a trace from another repetition), firing rate (middle) and raster plot of the motor units (bottom) followed by histogram of ISI normalized probability distribution of the extracted motor units in STIM01 for all repetitions during the maximum voluntary contraction task during SCS (40 Hz) at increasing amplitudes (7 mA, 10.8 mA, 12.5 mA; 2 repetitions for each stimulation amplitude). At 7 mA SCS, no motor units were detected. **b**, Histogram of ISI normalized probability distribution of the extracted motor units in STIM01 during 40 Hz SCS (above, 2 repetitions) and 60 Hz SCS (below, 2 repetitions) recorded in other sessions when performing the isometric task at 35% of maximum force. **c**, Histograms of ISI probability distribution of the extracted motor units in SCS04 and SCS03 with and without stimulation for all force levels for all trials (18 repetitions for SCS04 for all for levels, 9 repetitions in SCS03 for 5% isometric force level, 7 for 20%, 7 for 35% and 4 for 50%).

**Extended Data Fig. 9.**
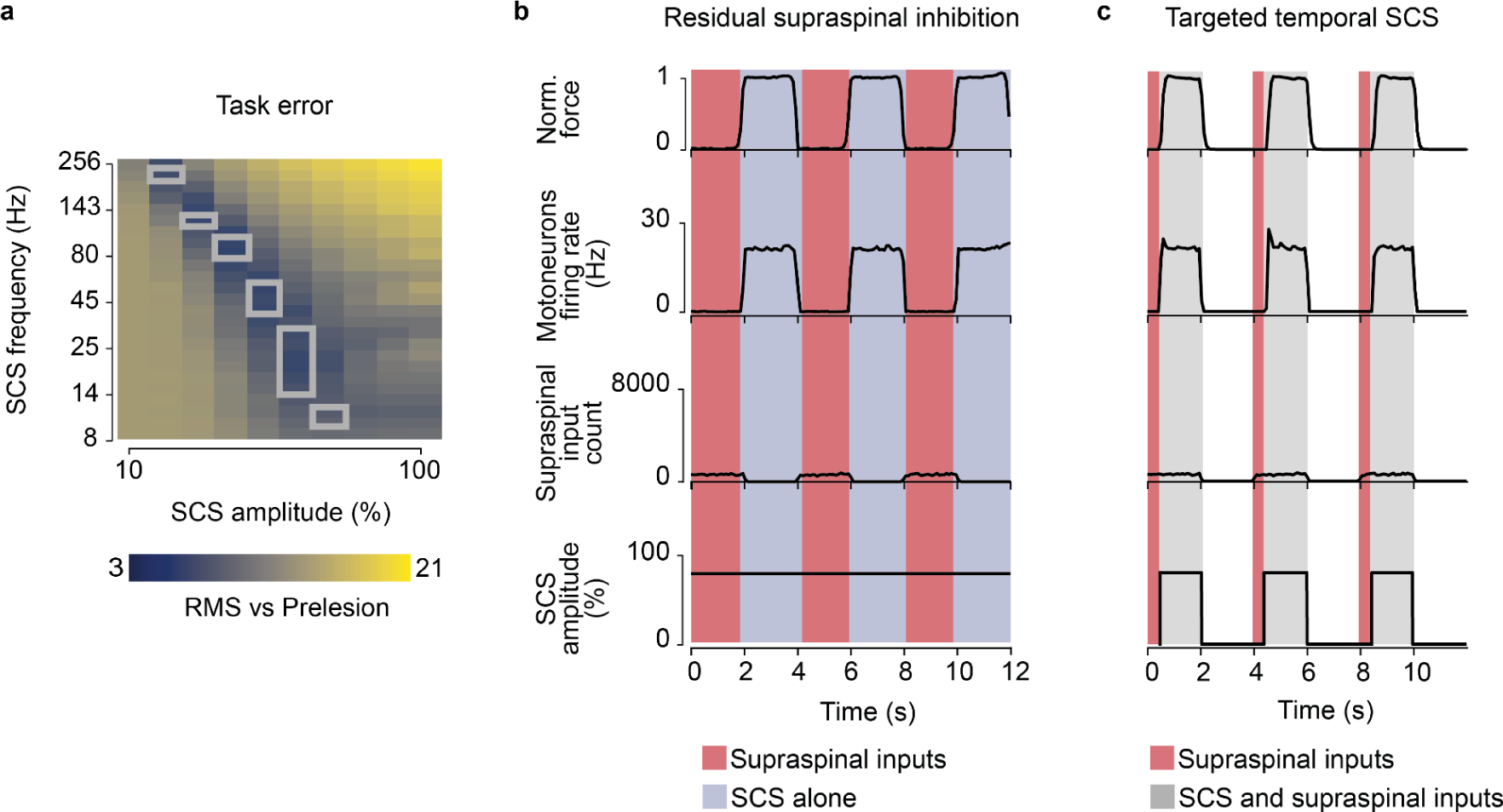
Simulation of SCS biophysics in a severe lesion. **a,** Root-mean square (RMS) of the difference between the motoneuron firing rate pre- vs postlesion for all simulated SCS parameters in a severe lesion (same as in **Fig. 4j**, but with a system with 11 supraspinal fibers). As with isometric force simulations in **Fig. 7b**, there is a shrinking of the voluntary movement regime where small task errors occur for a very restrained combination of SCS parameters. The gray lines in the heatmap indicate the voluntary movement regime for this severe lesion (**Fig. 7b**). **b,** Simulation of the normalized force and motoneuron pool firing rate during phasic activation of supraspinal fibers (2 seconds active, 2 seconds inactive) and continuous SCS at parameters belonging to the involuntary movement regime. Residual supraspinal fibers sculpt SCS drive ro produce functional motoneuron activity. Motoneurons firing is inhibited by the phasic activation of residual supraspinal fibers (red). **c,** Same as **b**, but SCS initiates 500 ms after the initiation of supraspinal fibers (red). Motoneurons now fire only coinciding with temporal activation of SCS.

**Extended Data Fig. 10.**
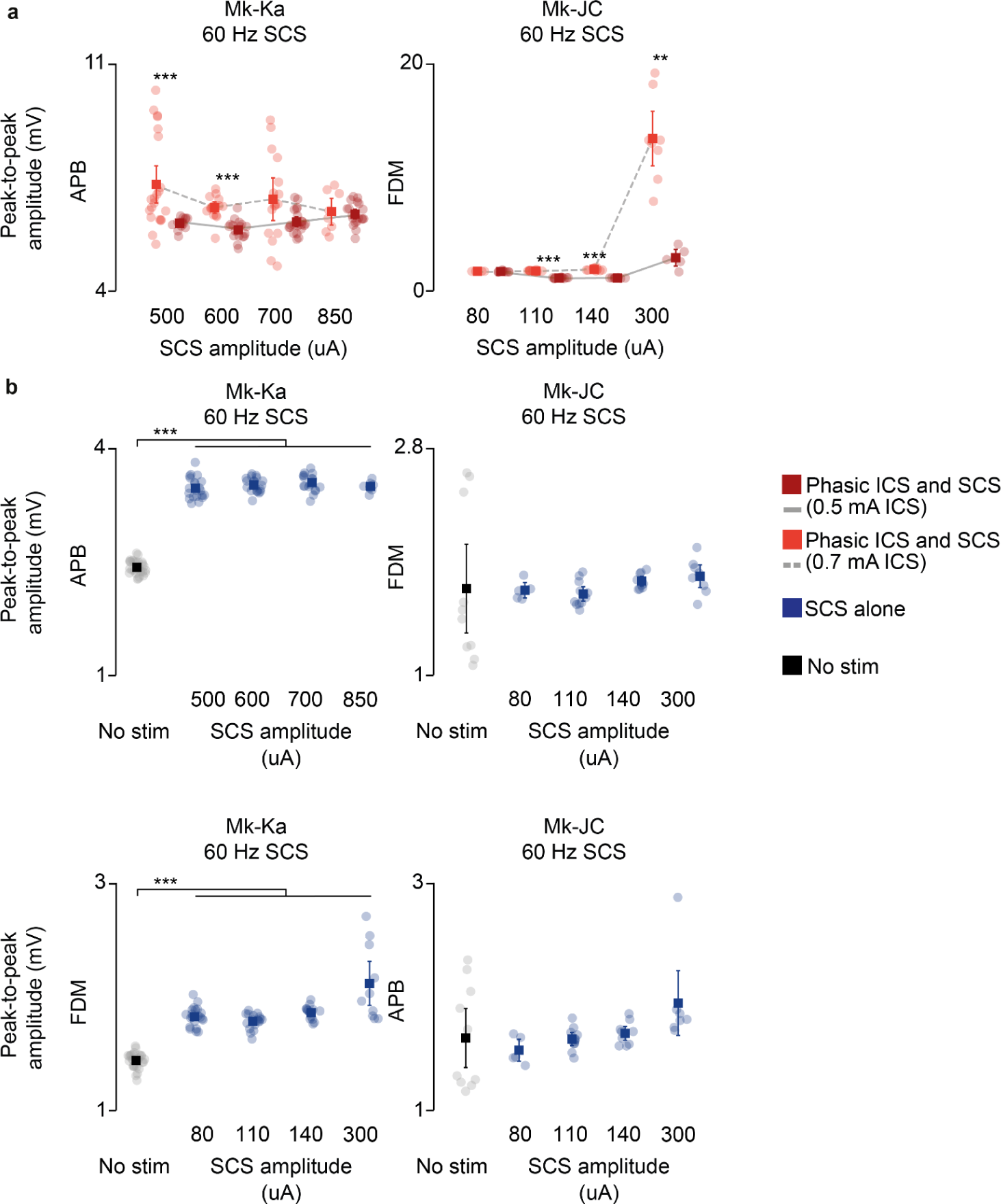
EMG responses during concurrent stimulation in monkeys. **a,** Root-mean square (RMS) of the EMG responses during phasic ICS and continuous SCS at increasing SCS amplitudes in addition to **Fig. 7e** (APB in Mk-Ka, FDM in Mk-JC). Statistical quantification of the RMS EMG responses (***p<0.001; **p<0.01; *p<0.05; Kruskal-Wallis test with 21, 14 points for 500 uA SCS at 0.7 mA ICS and at 0.5 mA ICS, respectively, in APB in Mk-Ka; 15, 18 points for 600 uA in Mk-Ka; 16, 21 points for 700 uA in Mk-Ka; 8, 19 points for 850 uA in Mk-Ka; 4, 8 points for 80 uA, 60 Hz SCS in Mk-JC; 11, 11 points for 110 uA, 60 Hz SCS in Mk-JC; 9, 8 points for 140 uA, 60 Hz SCS in Mk-JC; 8, 5 points for 300 uA, 60 Hz SCS in Mk-JC; 9, 10, 10 points for 80 uA, 30 Hz SCS in APB in Mk-JC; 8, 10, 8 points for 80 uA, 30 Hz SCS in FDM Mk-JC). **b**, RMS of the EMG responses during continuous SCS alone at increasing SCS amplitudes of the muscles shown in **a** (above) and **Fig. 7e** (below). Statistical quantification of the RMS EMG responses (***p<0.001; **p<0.01; *p<0.05; Kruskal-Wallis test with 27, 21, 19, 17 and 9 points for no stim, 500, 600, 700, 850 uA SCS, respectively, in APB in Mk-Ka; 10, 5, 11, 9, 8 points in FDM in Mk-JC; 27, 21, 18, 15, 10 points in FDM in Mk-Ka; 10, 5, 11, 9, 7 points in APB in Mk-JC). RMS EMG responses appeared to be significant in Mk-Ka, however, these responses were still below the lowest RMS value measured during concurrent phasic ICS.

## Notes

### Author Declarations

Institutional Review Board (IRB) of the University of Pittsburgh gave ethical approval for this work (protocols STUDY19090210 and STUDY22020031).

